# Metabolomic signatures associated with frailty, muscle strength and nutritional status in a cohort of older people

**DOI:** 10.1101/2025.05.14.25327600

**Authors:** Céline Bougel, Rémi Servien, Nathalie Vialaneix, Elise Maigne, Yves Boirie, Clément Lahaye, Cécile Canlet, Laurent Debrauwer, Valentin Max Vetter, Kristina Norman, Dominique Dardevet, Ilja Demuth, Sergio Polakof

## Abstract

**Background:** Frailty is a common geriatric syndrome characterized by increased vulnerability to stressors, reduced physiological reserves and heightened vulnerability and. It is also associated with adverse health outcomes. Given its complex phenotypes and underlying pathophysiology, there is a pressing need for robust, multidimensional biomarkers of frailty to advance personalized care. The objective of this study was to identify serum metabolomics signatures associated with different frailty phenotypes and related features.

**Methods:** We analyzed Nuclear Magnetic Resonance (NMR) metabolomics signatures in 901 subjects, 47.5% of them males (average age 68.34 ± 3.51 years) from the Berlin Aging Study II, categorized as non-frail, pre-frail, or frail based on Fried’s criteria at baseline (T0) and follow-up (T1, on average 7 years later). Linear models were used to assess associations between metabolite levels, frailty phenotypes, and frailty-related parameters.

**Results:** At baseline (T0), only 1% of the population was classified as frail, increasing to 4.8% at T1 (p-value<0.0001). In contrast, in terms of frailty progression during follow-up 323 subjects (35.8%) transitioned from non-frail to pre-frail, pre-frail to frail, or directly from non-frail to frail. In the overall population no significant differences were found in the relative quantifications of the 82 identified metabolites for any of the tested study outcomes. In contrast, 27 and 30 metabolites were negatively associated with handgrip strength at T0 and T1, respectively (*p*-value<0.05), and one metabolite (L-tyrosine, *p*-value=0.0297) positively associated with fat mass in men. In women, dimethylsulfone was positively associated with the percentage of evolution in the hand grip strength between T0 and T1 (*p*-value=0.0442), and glycerol was positively associated with appendicular lean mass at T0 (*p*-value=0.0049). Additionally, 22 metabolites were positively correlated with nutritional status at T0 in men (*p*-value<0.05): many of these were linked to carbohydrate (e.g., maltose, fructose, glucose, galactitol, mannose, lactate, acetylcarnitine) and amino acid metabolism (e.g., valine, tyrosine, isoleucine, alpha-hydroxybutyrate).

**Conclusions:** We may conclude that while serum metabolome revealed a weak association with frailty, significant associations were observed (particularly in men) between metabolomics signatures and frailty-related features such as muscle strength and nutritional status. These findings point to insulin sensitivity as a central feature, with early markers of impaired insulin sensitivity potentially impacting muscle health.

## Introduction

Frailty is a well-recognized geriatric phenotype ^1^ that signifies increased vulnerability and diminished physiological reserves ^2^. Its prevalence in the older population ranges from 4% to 60%, depending on the population studied. Frailty is characterized by a loss of physiological homeostasis and a reduced capacity to adapt to environmental changes ^3^, but also by impairment in various physiological processes and is associated with a number of adverse health outcomes including falls, morbidity, disability, hospitalization, institutionalization, and mortality ^4^.

Although there is broad consensus on the theoretical framework of frailty, its clinical identification remains challenging due to its complex pathophysiology, the heterogeneity of its phenotypic manifestations, and intra-individual fluctuations in severity. Additionally, while Fried’s Physical Frailty Phenotype (which comprises five components: unintentional weight loss, self-reported exhaustion, weakness, slow walking speed, and low physical activity) is widely adopted ^3^, the existence of multiple operational definitions further complicates diagnosis ^5^.

However, when comparing these assessment instruments, studies have found only modest overlap in their ability to identify frailty ^6^. This variability highlights the difficulty in detecting subtle deficits intrinsic to frailty, particularly biological and metabolic markers, which can impact the prediction of adverse outcomes. Two critical determinants of frailty — mobility and nutritional status—are especially important to capture. Nutrition plays a pivotal role in maintaining health and preventing frailty, as malnutrition or inadequate nutrient intake can lead to muscle wasting, reduced strength, and impaired function—key components of frailty. Conversely, frailty can exacerbate through poor appetite, eating difficulties, or nutrient deficiencies. Similarly, impaired mobility— manifested as reduced strength, balance issues, gait abnormalities, or falls—both signals and worsens frailty. Loss of mobility drives further muscle deconditioning and functional decline, reinforcing the progression of frailty.

A systematic review by Fernandez-Garrido et al. ^7^ emphasized the need to focus on the early stages of frailty development to better understand the mechanisms involved and their potential for reversibility. This would allow for timely and targeted healthcare interventions. Metabolomics has emerged as a powerful tool for molecular phenotyping, offering insights into the metabolic processes underlying various (patho-)physiological states and helping to identify biomarkers of metabolic dysregulation ^8^. These metabolomic biomarkers can detect early metabolic disturbances that precede the clinical manifestations of frailty, enabling healthcare providers to intervene in ways that could prevent or slow its progression, thereby reducing associated adverse outcomes. By analyzing the metabolomic profiles of frail individuals, researchers gain a deeper understanding of the biological mechanisms contributing to frailty, potentially paving the way to novel therapeutic strategies.

The objective of the present study was to better characterize the complexity of the frailty phenotype by determining the metabolomic signatures associated with frailty and its different individual components and related parameters, as well as their evolution over time. Based on data and serum samples from the Berlin Aging Study II (BASE-II) ^9^, we categorized participants as non-frail, pre-frail, or frail based on Fried’s criteria. The metabolomic profiles were analyzed at T0 (baseline) and T1 (follow-up, about 7 years later) using Nuclear Magnetic Resonance (NMR). We searched for associations between metabolite quantifications and frailty phenotypes as well as frailty associated features. Although we identified few associations with frailty, several components related to muscle strength or nutrition phenotypes yielded interesting results linked to early metabolic pathway degradation and insulin sensitivity.

## Materials and methods

### Study population and clinical data collection

In this study, older participants of the Berlin Aging Study II (BASE-II) ^9^ were analyzed. The BASE-II was conducted in accordance with the declaration of Helsinki and approved by the Ethics Committee of the Charité–Universitätsmedizin Berlin (approval number EA2/144/16 and EA2/144/16). All subjects gave written informed consent. The study is registered in the German Clinical Trials Register (Study-ID: DRKS00016157 and DRKS00016157).

A total of 1,083 subjects aged 60 years and older enrolled at baseline (T0, average age of 68.3±3.5 years, range 60.2-84.6 years, 2009-2014) completed medical assessments at follow-up (T1, average follow-up at 7.4±1.4 years, 2018-2020) (see the flowchart in **Figure 1**). Additionally, 17 participants were only examined at T1 resulting in a total sample size of 1,100 individuals. The medical assessment included parameters related to geriatric and internal medicine, along with phenotypic, nutritional, and functional data, collectively referred in this article as “clinical data” ^9^ to distinguish them from metabolomics data generated in the current project. For a detailed overview of the variables and categories, see **Supplementary Table 1**.

**Figure 1:**
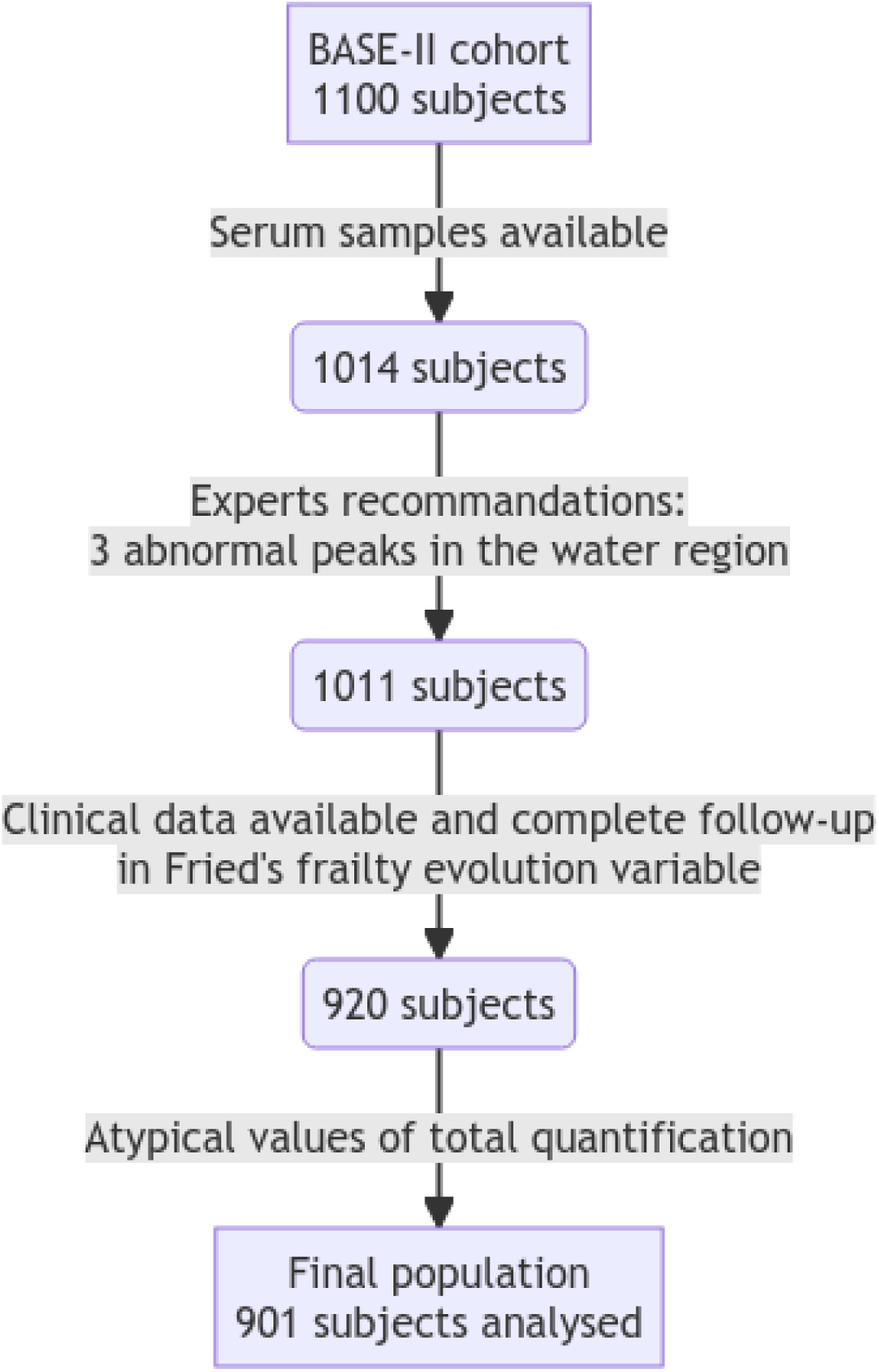
Flow chart of the study population for the present study, based on clinical and metabolomic data from subjects in the BASE-II cohort (n=1100 assessed at follow-up, T1). Three Nuclear Magnetic Resonance (NMR) samples were removed because they did not match the quality criteria. Then, 91 samples were removed due to missing data on the frailty evolution variable. Finally, 19 samples were removed due to extreme values in total quantification, resulting in poor overall reconstruction of the original spectrum by ASICS.

### Evaluation of the frailty status

Frailty was defined based on five criteria according to the definition proposed by Fried et al. ^3^, with some minor adjustments to align with the available BASE-II data ^10^. Pre-defined sex-specific cut-off values were used to award points in each category resulting in possible results between 0 and 5 points. Subjects were ranked as frail (3–5 points), pre-frail (1–2 points), or non-frail (0 point). Unintentional weight loss (one point) was defined as losing at least 5% of body weight in the past year. The self-reported exhaustion was determined based on two questions from the Center for Epidemiological Studies depression scale (CES-D ^11^): exhausted (1 point) or not exhausted (0 point). Weakness was assessed by measuring hand grip strength using sex-and BMI-specific thresholds ^3,10^. Those who fell below the threshold were considered weak and received one point. Walking speed was evaluated with the “Timed Up & Go” test ^12^: participants taking more than 10 seconds to complete the test were classified as having slow walking speed. Low physical activity was determined by asking, “Are you seldom or never physically active?”. Those who answered “Yes” were considered physically inactive. Each of the previously mentioned criteria, when met, contributed one point to the frailty score ^10^.

Statistical analyses were also performed on each component of the Fried’s frailty score, as a categorical index (yes/no) or as a numeric variable (whenever available). In the latter case, the percentage of evolution between the baseline and the follow-up and absolute change over the same period were analyzed similarly to the raw data. As done for previous analyses in BASE-II ^10,13^, participants in the “frail” and “pre-frail” group were combined and referred to as “pre-frail + frail” in all analyses.

In addition, a variable for the evolution of frailty within the on average 7.4±1.5 years between baseline and follow-up assessments was created, based on Fried’s frailty index categories, as follows (**Figure 2**): “controls” for non-frail subjects throughout the follow-up; “stable” for those who had the same level of frailty (other than non-frail) throughout the follow-up; “improve” for frailty enhancement (e.g. frail to pre-frail), and “worsened” for frailty degradation (e.g. non-frail to pre-frail).

**Figure 2:**
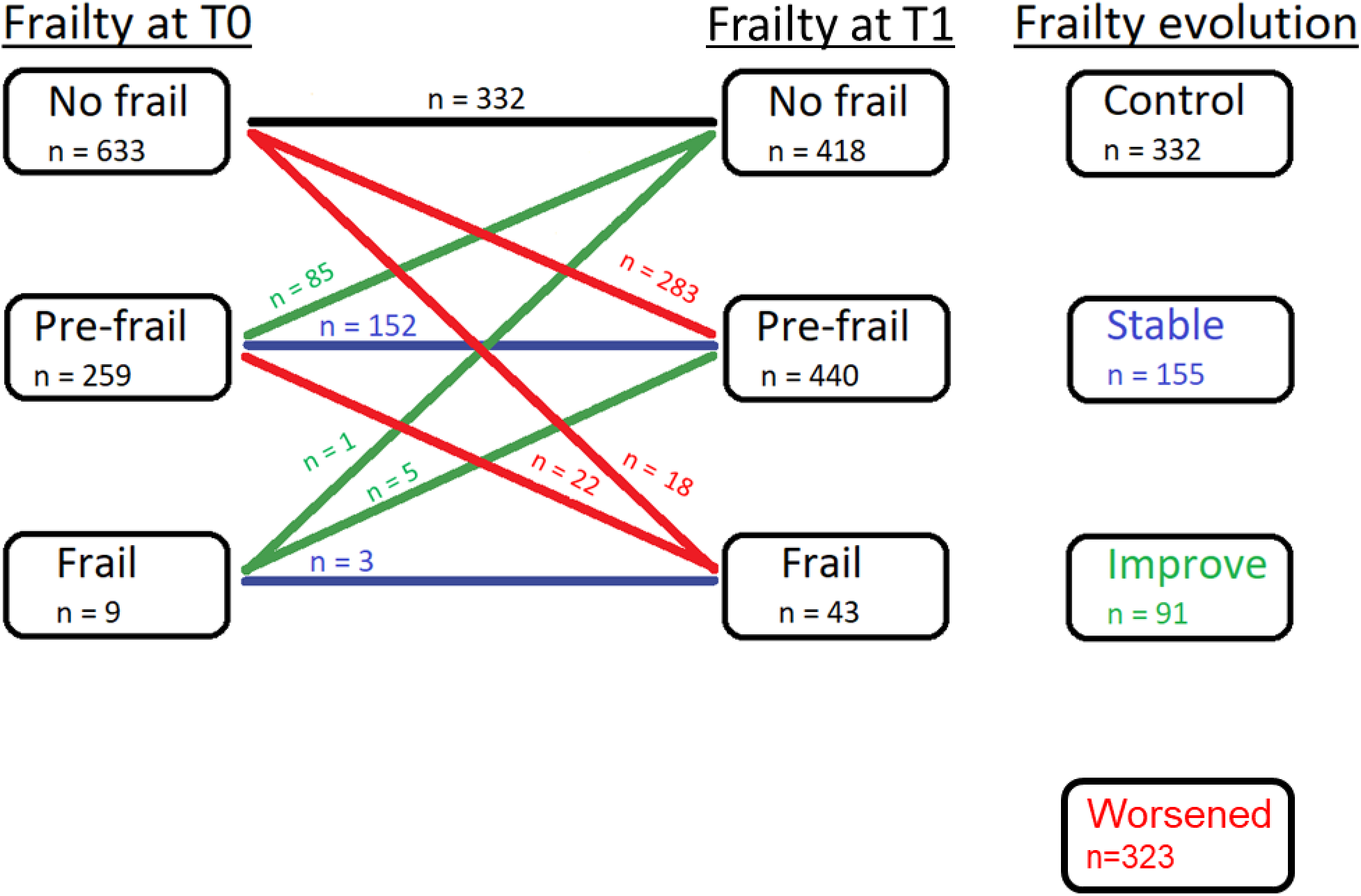
Study design and frailty status evolution of the 901 subjects over the 7-year period, using Fried’s frailty index. Four sub-groups of subjects were defined: Control subjects who remain no frail over time [from non-frail at the recruitment to non-frail at the end of follow-up]; Stable subjects who did not change their frailty status over time but with a different level from control subjects [from pre-frail to pre-frail and from frail to frail]; those who improved their frailty status [from pre-frail to non-frail, from frail to pre-frail and from frail to non-frail] and those who worsened their frailty status [from non-frail to pre-frail, from pre-frail to frail and from non-frail to frail] over follow-up.

### Frailty-related parameters

We have also evaluated other variables related to frailty, including appendicular lean mass (ALM, sum of lean tissue in the arms and legs in kg), body fat mass (kg/m²) from DXA scans, and several variables related to the nutritional status of the participants: total dietary leucine intake (g/day), total protein intake (g/day), total daily energy intake assessed via food frequency questionnaire, and overall nutritional status (estimated using the Mini Nutritional Assessment (MNA^14^).

### ^1^H NMR data acquisition: analyses of serum samples, data processing, and metabolite identification and quantification

Serum samples (100 µL) were mixed with 200 µL of phosphate buffer (pH 7.0). After centrifugation (5500 g, 4°C, 15 min), 200 µL of supernatant were transferred into 3 mm NMR tubes. ^1^H NMR spectra were obtained at 300 K on a Bruker Avance III HD 600 MHz NMR spectrometer (Bruker Biospin, Rheinstetten, Germany), operating at 600.13 MHz for ^1^H resonance frequency using an inverse detection 5 mm ^1^H-^13^C-^15^N-^31^P cryoprobe attached to a Cryoplatform (the preamplifier unit). “Tuning” and “matching” of the probe, lock, shims tuning, pulse (90°) and gain computation were automatically performed for each sample. ^1^H NMR spectra were acquired using the 1D CPMG experiment for suppression of macromolecule signals and presaturation for water suppression. A total of 128 transients was collected into 32k data points using a spectral width of 20 ppm, a relaxation delay of 2 s, and an acquisition time of 2.72 s. Before the Fourier transform, an exponential line broadening function of 0.3 Hz was applied to the FID. All NMR spectra were phase-and baseline-corrected and referenced to the chemical shift of TSP (0 ppm) using Topspin (V3.2, Bruker Biospin, Germany).

NMR spectra were then divided into fixed-size buckets (0.01 ppm) between 9.0 and 0.5 ppm using the AMIX software (v3.9.15, Bruker), and area under the curve was calculated for each bucket (integration). The region including residual water (5.1-4.5 ppm) was removed. Buckets were normalized according to the total intensity. Preprocessed data were then exported into a “1r format” file for data treatment and statistical analysis.

^1^H NMR spectra were processed using the R package ASICS, version 2.10 ^15^ with the R software version 4.1.3 (2022-03-10) ^16^. Standard preprocessing of the spectra (as described in ^15^, including 1r format signals importation, water area removal, and baseline correction were performed. All the 920 spectra (see the flowchart in **Figure 1**) were then aligned with each other using the ASICS joint alignment procedure ^17^.

Automatic quantifications of metabolites were performed using the ASICS joint quantification procedure and options quantif.method = “both” and clean.thres = 0.25 as advised in ^17^. The max.shift, noise.thres, add.noise, and mult.noise parameters were set to their default values. It is important to note that the metabolite quantifications presented here are relative quantifications (see ^15^ for details).

### Statistical analyses

All statistical analyses were carried out using R ^16^. After quality control analyses of the 1,014 participants initially available, a total of 901 eventually has a complete dataset with no missing values in metabolomics or frailty score measures (see **Supplementary methods** for more details).

Linear regression models were fitted to assess statistical associations between metabolite quantifications and study outcomes (including frailty, individual frailty components, and frailty-related variables). Since sex differences are known to play a role in all major diseases, their prevention and treatment and since aging also interacts with sex-related health differences ^9^, a linear regression model was first used to address the following question: “Which metabolites exhibit a significant interaction effect between study outcomes and sex?”:

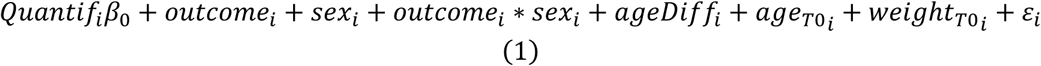

against the alternative model (Fisher test)

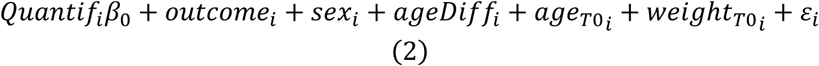

where:

- Quantif*_i_* is the vector of the quantification of the tested metabolite for subject *i,*
- β_0_ is the intercept, and ε*_i_* is the error of the model, ε*_i_* ∼ N(0, σ^2^),
- sex*_i_* is the subject sex (man/woman),
- study outcome*_i_* is one of the variables of interest, including frailty, frailty individual components and frailty-related parameters, at baseline and at follow-up,

Three numeric covariates were included in the model: ageDiff*_i_* is the duration of follow-up (difference between age at T1 and age at T0), age_T0*_i_* is the age at T0, and weight_T0*_i_* is the body weight at T0.

Given that the sex effect was found important (**Supplementary Figure 3**), we decided to fit other linear models in the two sex populations:

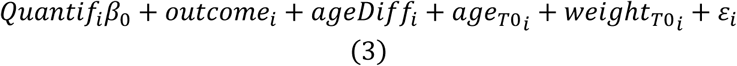

against the alternative model

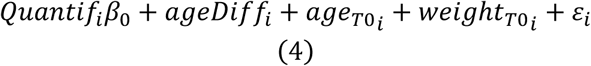

(same notations as above, Fisher test).

When looking at these variables stratified by sex, a bimodality in the distribution of the duration of the follow-up variable emerged (see Supplementary Figure 5). This discrepancy is due to the study’s enrollment process: initially, a first wave of recruitment included a higher proportion of men, followed by a second wave that balanced the sex distribution. Although the study procedures were the same for all participants, the difference in mean follow-up time (subjects were followed for an average of 7.43 ± 1.43 years) could influence the analysis results. Therefore, follow-up time was included as a control covariate in the models.

For a given study outcome, *p*-values were adjusted for multiple testing using the Benjamini and Hochberg (BH) procedure to control the False Discovery Rate. Metabolites with adjusted *p-* values below 5% were considered as significant, while *p*-values between 5-10% are referred as “trend association”.

Pathway enrichment analysis

Metabolic pathway enrichment analysis was performed using the MetaboAnalyst online web interface (https://www.metaboanalyst.ca/, version 5.0), which supports metabolic pathway analysis (integrating pathway enrichment analysis with a hypergeometric test, and pathway topology analysis). The reference metabolome included in the analysis contained the metabolites present in the ASICS pure spectra library. The results of the various models, corresponding to the previously presented study outcomes, were analyzed separately.

## Results

### BASE-II participants were overall robust

Clinical data for the 901 subjects in the present study are summarized in **Table 1 and 2**. The average age was 68.34±3.51 years at T0 and 75.77±3.81 years at T1. The number of participants was balanced according to sex: 428 men (47.5%) and 473 women (52.5%). Their average weight at T0 was 76.83±14.08 kg, with only a minor reduction of 1.08%±6.47 (*p*-value<0.0001, paired Student’s *t*-test, **Table 1**), as previously observed by Vetter et al. ^18^.

**Table 1:** Summary of numerical variables for the main parameters explored in the present study.

| Variable | Mean±SD<br>at T0 | Mean±SD<br>at T1 | P-value* | Percentage of<br>evolution T0-T1 |
| --- | --- | --- | --- | --- |
| Age (years) | 68.34±3.51<br>(n=901) | 75.77±3.81<br>(n=901) | <0.0001 | 10.90±2.15<br>(n=901) |
| Weight loss (kg) | 76.83±14.08<br>(n=899) | 75.88±13.96<br>(n=901) | <0.0001 | 1.08±6.47<br>(n=899) |
| Body Mass Index (kg/m<sup>2</sup>) | 26.83±4.11<br>(n=901) | 27.78±3.81<br>(n=887) | 0.0045 | 0.81±6.61<br>(n=887) |
| ALM<sup>1</sup> (kg) | 21.22±4.98<br>(n=863) | 20.28±4.73<br>(n=546) | <0.0001 | 2.83±7.31<br>(n=543) |
| Fat mass (kg/m<sup>2</sup>) | 9.15±2.64<br>(n=475) | 9.25±2.53<br>(n=540) | 0.0906 | 1.98±14.19<br>(n=471) |
| CES-D<sup>2</sup> (points) | 13.90±3.52<br>(n=889) | 13.58±3.74<br>(n=896) | 0.0270 | 2.20±35.39<br>(n=884) |
| Hand grip strength<sup>3</sup> (kg) | 34.15±9.70<br>(n=901) | 27.30±9.16<br>(n=901) | <0.0001 | 20.08±14.35<br>(n=901) |
| RAPA<sup>4</sup> (points) | 5.21±1.45<br>(n=898) | 4.78±1.25<br>(n=901) | <0.0001 | 1.70±53.21<br>(n=898) |

| Variable | Mean±SD<br>at T0 | Mean±SD<br>at T1 | *P*-value* | Percentage of<br>evolution T0-T1 |
| --- | --- | --- | --- | --- |
| Leucine intake (g/day) | 5.99±2.08<br>(n=884) | 5.73±2.11<br>(n=859) | <0.0001 | 0.88±28.67<br>(n=843) |
| Protein intake (g/kg/day) | 1.06±0.36<br>(n=882) | 1.02±0.35<br>(n=859) | 0.0015 | 0.41±29.64<br>(n=843) |
| Protein intake (g/day) | 79.98±28.13<br>(n=884) | 76.38±28.35<br>(n=859) | <0.0001 | 1.01±28.61<br>(n=841) |
| Total energy intake (kJ/day) | 9,408±2,957<br>(n=884) | 9,192±3,068<br>(n=859) | 0.0382 | 1.18±29.16<br>(n=843) |
| MNA<sup>5</sup> score (points) | 27.12±1.80<br>(n=878) | 26.32±2.20<br>(n=876) | <0.0001 | 2.71±9.00<br>(n=854) |

\* *P*-value of the paired Student's *t*-test for average comparison between T0 and T1, i.e. the test is only performed on subjects who can be matched; # NA for non-applicable;
<sup>1</sup> Appendicular lean mass (ALM); <sup>2</sup> Center for Epidemiological Studies depression scale (CES-D); <sup>3</sup> Rapid Assessment of Physical Activity (RAPA) score.
<sup>4</sup> sex- and BMI-specific thresholds applied
<sup>5</sup> Mini Nutritional Assessment (MNA); Data is presented as mean Mean±SD or percentage (number of subjects).

As already reported by Vetter et al.^13^, at T0, only a small proportion of subjects were considered as frail according to Fried’s frailty index (1.0%) (see **Table 2**), while this proportion significantly increased to 4.8% at follow-up (*p*-value<0.0001, McNemar’s chi-square test, in **Table 2**). The study population was therefore predominantly robust at baseline according to Fried’s frailty index, with 892 subjects classified as non-frail or pre-frail (633 and 259 respectively). In terms of frailty progression during follow-up (see **Table 2**), 323 subjects (35.8%) were categorized in the ‘worsened’ group, meaning they transitioned from non-frail to pre-frail, pre-frail to frail, or directly from non-frail to frail. Interestingly, 91 subjects (10.1%) showed an improvement in frailty status, transitioning from pre-frail to non-frail, frail to pre-frail, or frail to non-frail according to Fried’s frailty index. Concerning the individual components of Fried (**Table 3**), we observed a slight increase in the proportion of subjects with an exhaustion perception, reaching 11.3% at the T1 follow-up (*p*-value=0.0112, McNemar’s chi-square test). In addition, the proportion of subjects classified as weak importantly increased (5-fold) at the follow-up, reaching 30.5% (*p*-value<0.0001, McNemar’s chi-square test). Similarly, the number of subjects with slow walking speed doubled over the follow-up from 10.7% to 20.9% (*p*-value<0.0001, McNemar’s chi-square test). Finally, while the physical inactivity score was slightly reduced from 5.21 points at T0 to 4.78 points at T1 (*p*-value<0.0018, McNemar’s chi-square *t*-test): 87.8% of the population was considered physically active at the follow-up examination.

**Table 2:** Summary of categorical variables for the main study outcomes of the present study.

| Variable | Modalities | Number of subjects<br>(%) at T0 | Number of subjects<br>(%) at T1 | P-value<sup>1</sup> |
| --- | --- | --- | --- | --- |
| *Sex* | Man | 428 (47.5) | Not Applicable | Not Applicable |
|  | Woman | 473 (52.5) | Not Applicable |  |
| *Fried frailty evolution* | Control | 332 (36.8) | Not Applicable | Not Applicable |
|  | Stable | 155 (17.2) | Not Applicable |  |
|  | Improve | 91 (10.1) | Not Applicable |  |
|  | Worsen | 323 (35.8) | Not Applicable |  |
| *Fried frailty index* | No frail | 633 (70.3) | 418 (46.4) | <0.0001 |
|  | Pre-frail | 259 (28.7) | 440 (48.8) |  |
|  | Frail | 9 (1.0) | 43 (4.8) |  |
| *MNA<sup>3</sup> index* | Normal | 852 (94.6) | 776 (86.1) | <0.0001<sup>2</sup> |
|  | Malnourished | 26 (2.9) | 100 (11.1) |  |
|  | Missing | 23 (2.5) | 25 (2.8) |  |

<sup>1</sup> P-value of the McNemar's chi-square test proportion comparison between T0 and T1.
<sup>2</sup> P-value of the corrected McNemar's chi-square test proportion comparison between T0 and T1.
<sup>3</sup> Mini Nutritional Assessment (MNA).

**Table 3:** Summary of the five categorical components of Fried’s frailty index.

| Variable | Modalities | Number of subjects<br>(Percentage)<br>at T0 | Number of<br>subjects<br>(Percentage) at T1 | *P*-value<sup>1</sup> |
| --- | --- | --- | --- | --- |
| Unintentional weight loss |  |  |  | 0.5677<sup>2</sup> |
|  | No | 877 (97.3) | 872 (96.8) |  |
|  | Yes | 24 (2.7) | 29 (3.2) |  |
| Exhaustion |  |  |  | 0.0112 |
|  | Not exhausted | 827 (91.8) | 799 (88.7) |  |
|  | Exhausted | 74 (8.2) | 102 (11.3) |  |
| Weakness |  |  |  | <0.0001 |
|  | No weak | 842 (93.5) | 626 (69.5) |  |
|  | Weak | 59 (6.5) | 275 (30.5) |  |
| Walking speed |  |  |  | <0.0001 |
|  | Not slow | 805 (89.3) | 713 (79.1) |  |
|  | Slow | 96 (10.7) | 188 (20.9) |  |
| Physical activity |  |  |  | 0.0018 |
|  | Not low | 829 (92.0) | 791 (87.8) |  |
|  | Low | 72 (8.0) | 110 (12.2) |  |

<sup>1</sup> *P*-value of the McNemar's chi-square test proportion comparison between T0 and T1.
<sup>2</sup> *P*-value of the corrected McNemar's chi-square test proportion comparison between T0 and T1.

Regarding other numerical variables related to body composition and muscle health (**Table 1**), appendicular lean mass averaged 21.22±4.98 kg at T0 and significantly decreased to 20.28±4.73 kg at T1 (*p*-value<0.0001, paired Student’s *t*-test). This was accompanied by a similar reduction in hand grip strength averaged 34.15±9.70 kg at T0, and subjects lost 20.08%±14.35 of their initial grip strength 7 years later (*p*-value<0.0001, paired Student’s *t*-test). In contrast a more modest reduction (1.7%, *p*-value<0.0001, paired Student’s *t*-test) in the declared physical activity was observed (RAPA test). Finally, the subjects’ fat mass (9.15±2.64 kg/m² at T0), did not increase significantly over seven years (*p*-value=0.09, paired Student’s *t*-test).

Concerning nutrition-related variables (**Table 1**), only minor changes were observed after the 7 years follow-up, including-1.18%±29.16 for total energy intake (*p*-value=0.0382, paired Student’s *t*-test),-0.88%±28.67 for leucine intake (*p*-value<0.0001, paired Student’s *t*-test) and-0.41%±29.64 for protein intake (*p*-value<0.0001, paired Student’s *t*-test). The MNA identified only 26 malnourished subjects at T0 (**Table 2**), while the number increased by 3-fold at the follow-up (*p*-value<0.0001, McNemar’s chi-square test with continuity correction, **Table 2**), reaching 100 individuals.

### Metabolomics signatures were poorly associated with frailty, but rather with frailty-related phenotypes

Metabolites with significantly different quantifications according to individual frailty components or frailty-related parameters in each population (overall, men and women, respectively), are summarized in **Table 4**, while average quantifications and *p*-values can be found in **Supplementary Tables 2and 3** respectively.

**Table 4:**
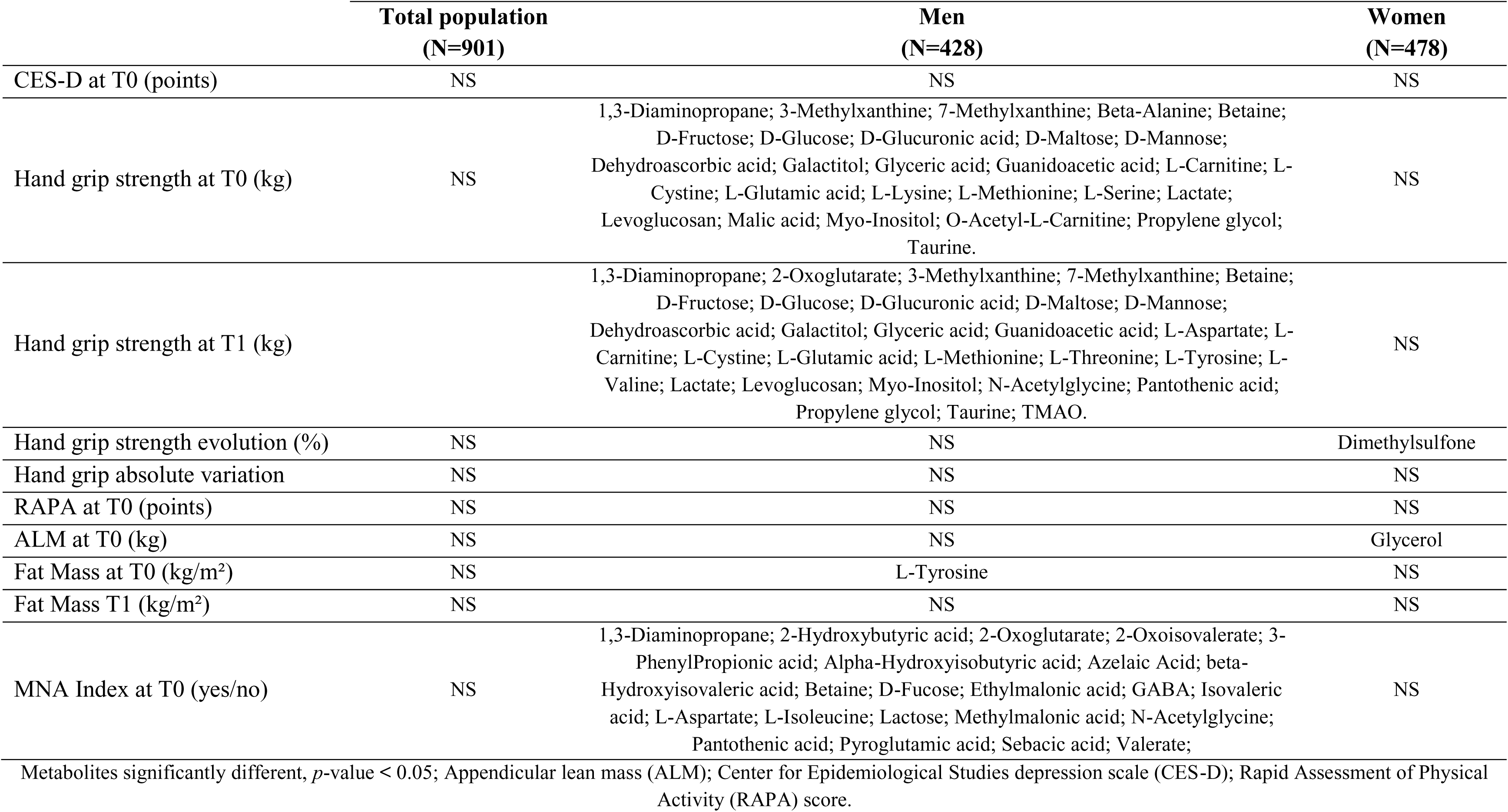
Summary of metabolites positively associated with individual frailty components or frailty-related parameters, according to subject sex, based on linear Models (3) and (4). N=901 subjects of the BASE-II cohort.

In the overall population no significant differences were found in the relative quantifications of the 82 identified metabolites for any of the tested study outcomes. Concerning the frailty-related phenotypes, 37 metabolites showed trend associations (i.e. *p*-value between 5% and 10%) with the hand grip strength at T1 and dimethylsulfone showed trend association with the absolute variation of the hand grip strength between T0 and T1 (**Supplementary Table 3)**. In women, dimethylsulfone was positively associated with the percentage of evolution in the hand grip strength between T0 and T1 (adjusted *p*-value=0.0442) (**Figure 3**). In contrast to women, numerous metabolites were associated with several frailty-related items in men (**Table 4 and Figure 4 and 5**). We found significant negative associations between hand grip strength and several quantified metabolites: 27 at T0, and 30 metabolites at T1. Many other additional metabolites showed also trend associations, including 12 metabolites at T0 and 18 metabolites at T1 with the hand grip strength, dimethylsulfone with the self-reported exhaustion at T0, and 14 metabolites with physical inactivity score at T0 (**Supplementary Table 3**).

**Figure 3:**
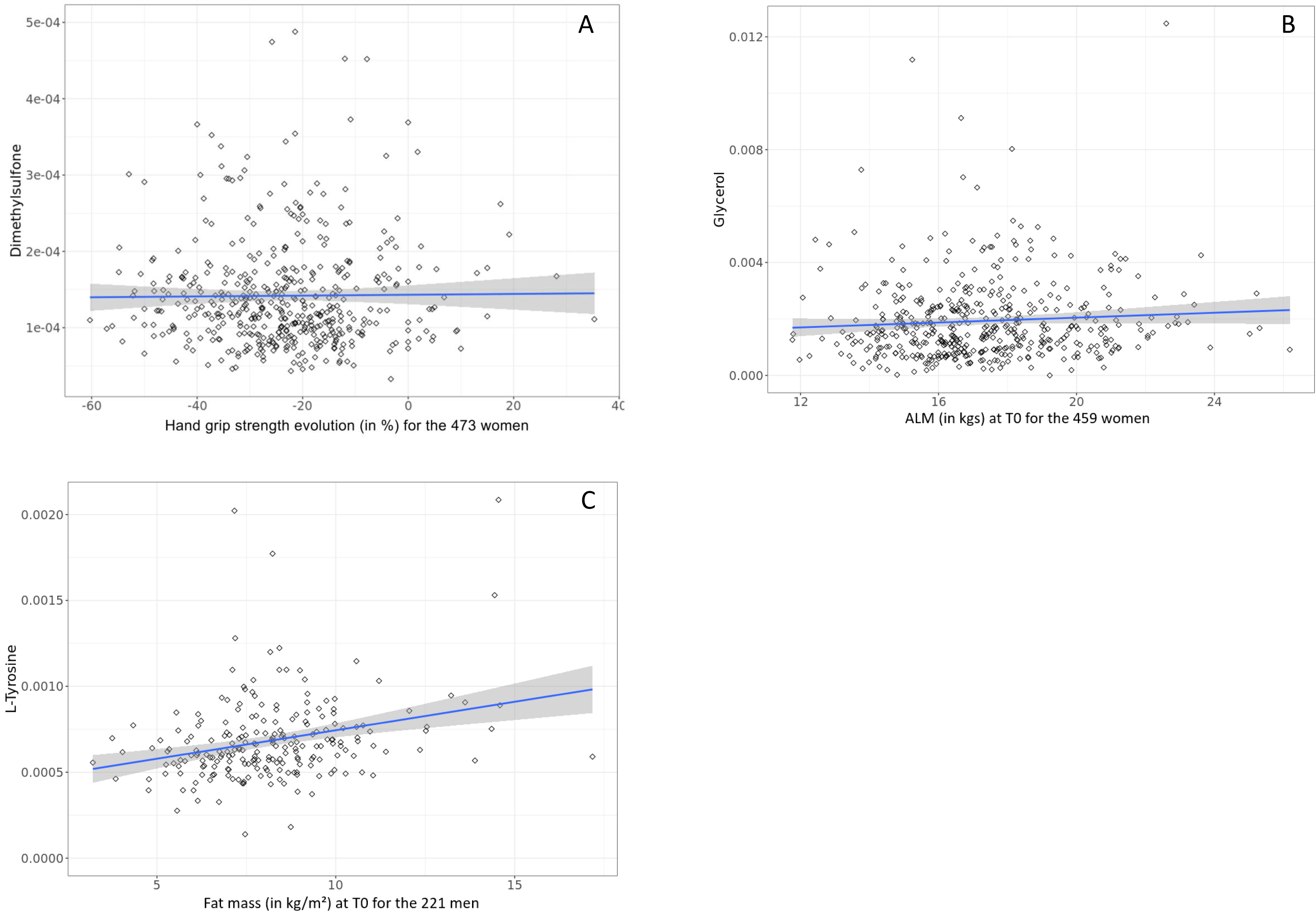
Scatterplot of significant metabolites in the linear model of the second research question (Models (3) and (4)). For frailty-related parameters, Dimethylsulfone was associated with the percentage of evolution of the hand grip strength at T0 (A) for women of the BASE-II cohort (two subjects with quantification > 0.0005 were hidden for better readability). Glycerol was associated with the appendicular lean mass (ALM) at T0 (B) for the women; and L-Tyrosine as a function of body fat masse at T0 (C) for the men. The number of subjects represented on the graphs, and their sex is indicated in the x-axis label for each variable.

**Figure 4:**
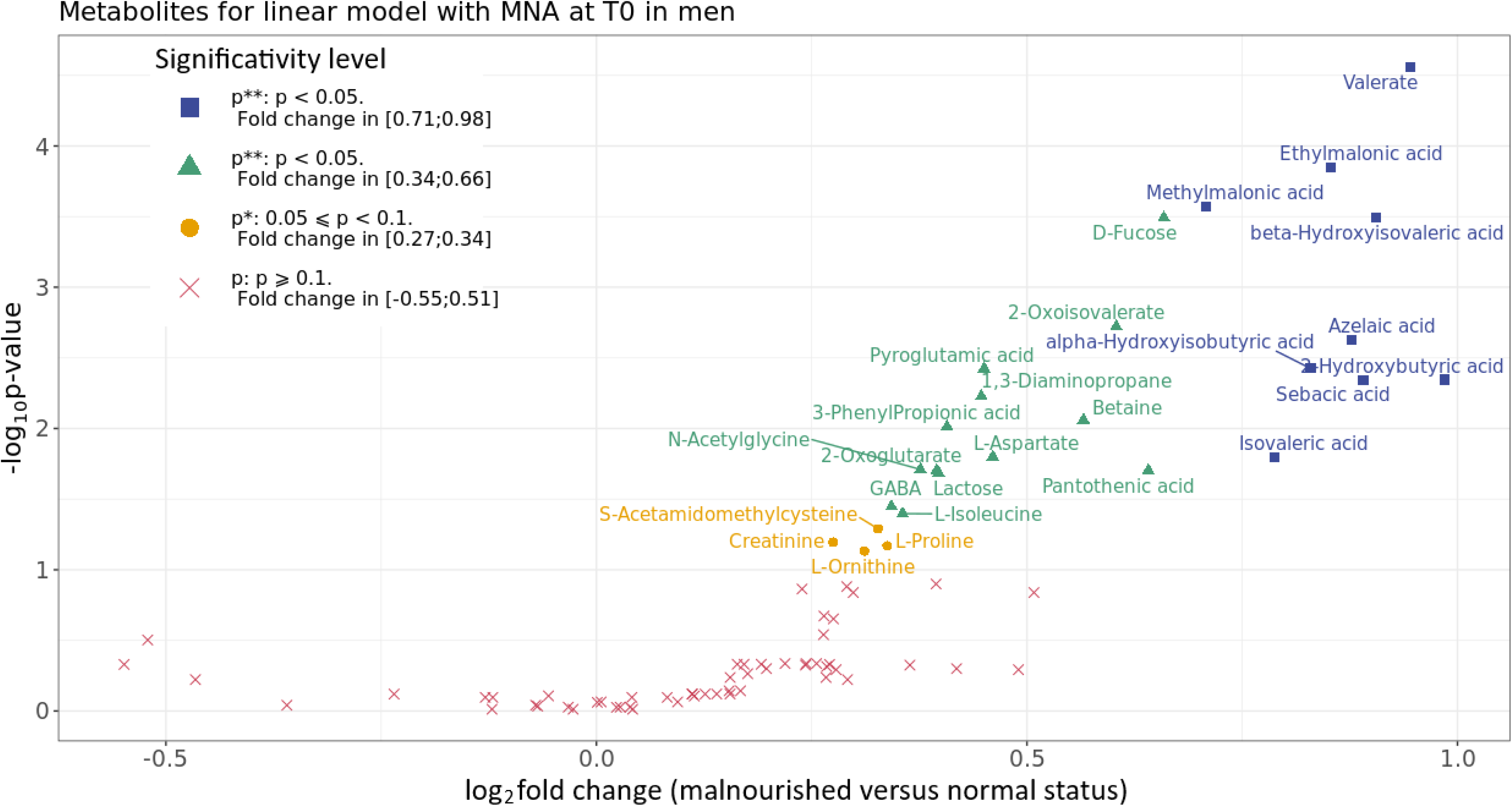
Volcano plot of metabolomic data, for differential metabolites in men of the BASE-II cohort (N=428). The y-axis is the negative logarithm in base 10 of the adjusted *p*-values obtained from the linear model of the second research question (Models (3) and (4) of Section 2.4.2, for the Mini Nutritional Assessment (MNA) index at baseline). The x-axis is the metabolite quantification fold change (plotted on a logarithm in base 2) as a ratio between the two nutritional status (malnourished versus normal status). The log2 Fold Change indicates the average quantification level for each metabolite. Each dot represents a different metabolite. The colors and shapes were associated with significance and expression levels: red cross for unchanged expression and non-significant metabolites (i.e. adjusted *p*-value > 0.1), yellow dots for unchanged expression and tend to significant metabolites (i.e. adjusted *p*-value between 0.05 and 0.1), green triangle for unchanged expression and significant metabolites (i.e. adjusted *p*-value below 0.05), and blue square for up-regulated and significant metabolites (i.e. adjusted *p*-value below 0.05).

**Figure 5:**
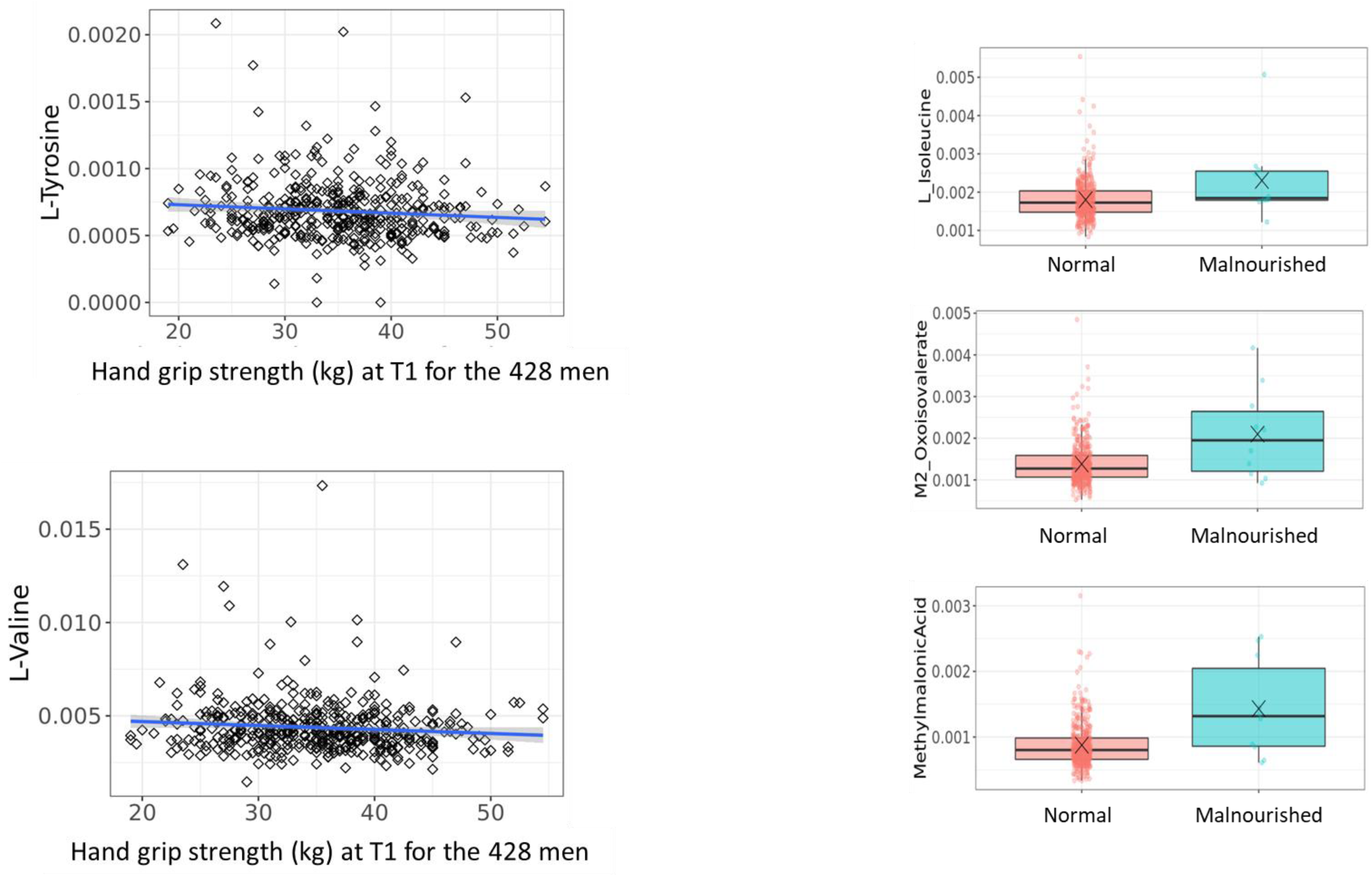
Selected metabolites related to the BCAA/AAA metabolism and identified as associated to muscle strength and/or the nutritional status (MNA) in the BASE II cohort. More details in Tables 5 and 6.

**Table 5:**
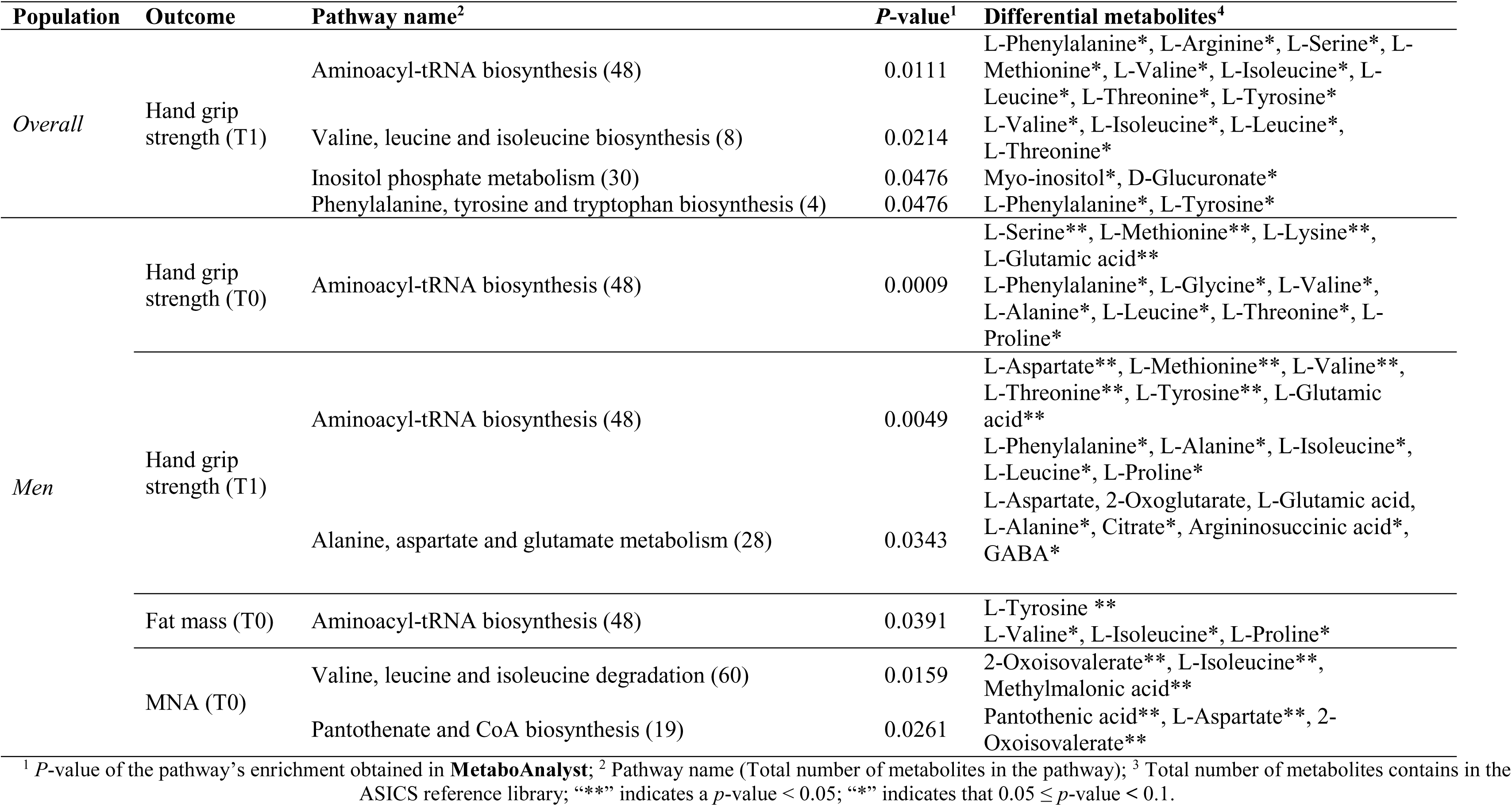
Enriched pathways in differential metabolites with an adjusted p-value below 10% for linear models.

Regarding those body composition and muscle health, glycerol was positively associated with appendicular lean mass at T0 in women (adjusted *p*-value=0.0049) and L-Tyrosine was positively associated with body fat mass at T0 in men (adjusted *p*-value=0.0297) (**Figure 3**). TMAO showed a trend association with body fat mass at follow-up in the whole population, while 13 additional metabolites (including 3 AA, namely isoleucine, proline and valine) showed a trend association in men (**Supplementary Table 3**).

Concerning the nutritional-related parameters, analyses revealed changes in the relative quantifications only for men: 22 metabolites were positively associated with MNA score at T0 (**Table 4**), i.e. the relative levels were higher in malnourished subjects when compared to subjects with a normal nutritional status (**Figures 4 and 5**), while four others (Creatinine, L-Ornithine, L-Proline, S-Acetamidomethylcysteine) showed a trend (**Supplementary Table 3**). Interestingly, 22 metabolites were significantly associated with the hand grip strength at T0 and at T1. Five of them were specific to the association with the hand grip strength at T0 (Beta-Alanine, L-Lysine, L-Serine, Malic Acid, and O-Acetyl L-Carnitine) and eight of them were specific to the association with the hand grip strength at T1 (L-Aspartate, L-Threonine, L-Tyrosine, L-Valine, 2-Oxoglutarate, N-Acetyl glycine, Pantothenic Acid, and TMAO). **Table 5** summarized the metabolic pathway enrichment analyses, for significant metabolites or trend associations described above.

## Discussion

Frailty is a multifaceted condition that manifests differently across individuals in the general population. Investigating its early stages, such as pre-frailty, is crucial for both prevention and a deeper understanding of the condition’s underlying mechanisms ^7^. In this study, although the NMR-based metabolomics signatures were not significantly associated with frailty (as defined by the Fried criteria), numerous metabolites were associated with two key frailty-related factors, namely muscle strength and nutrition-associated variables. Many of these metabolites, including several sugars, amino acids, and others, were linked to a common underlying factor: insulin sensitivity.

### Frailty is weakly associated with the serum NMR metabolome in BASE-II participants

Our results indicate that, when examining both men and women from the BASE-II population, frailty does not manifest in the metabolomic profile. This was also true for the individual categorical components of frailty, including body weight loss, gait speed, fatigue, physical activity, and muscle weakness. These findings align with recent studies conducted on older Chinese and Italian populations, where either frailty or its components were only moderately distinguished ^8,19^, and changes in specific metabolites, primarily lipids, were deemed modest in various case-control designs^20^. In our study, the weak signal of frailty within the metabolome may be attributed to the low number of frail individuals (9 at baseline and 43 at follow-up) in the cohort. Additionally, definitions of frailty differ and it is possible that the definition by Fried covers aspects of frailty that are not strongly connected to the metabolome ^21^.

### Reduced muscle strength could be associated with dysregulated metabolism at the muscle level

We found that numerous metabolites in men were negatively associated with muscle strength at both baseline and follow-up. Notably, energy metabolism appeared to play a crucial role in these associations. Carbohydrate metabolism was central to the metabolomic signature associated with muscle strength, involving six metabolites (see **Table 4**), most of which are upstream in the carbohydrate oxidation pathway (maltose, fructose, glucose, galactitol, mannose) and one is downstream (lactate). Carbohydrates serve as the primary energy source for muscle activity, efficiently oxidized through glycolysis to generate ATP during physical exertion, while also contributing to glycogen replenishment afterwards. Additionally, carbohydrates play a vital role in the protein-sparing effect. Insufficient or dysregulated carbohydrate supply for energy production can lead to increased muscle protein breakdown to oxidize amino acids, thereby contributing to muscle wasting. The accumulation of these metabolites in circulation may result from impaired mitochondrial function observed in frailty, as well as insulin resistance, as illustrated by the muscle atrophy reported in Type 2 Diabetes (T2D) patients ^22^. This is further supported by the negative correlation between muscle strength and other metabolites related to reduced insulin sensitivity, such as acetylcarnitine, valine, and tyrosine (**Figure 5**). Acetylcarnitine, the shortest acylcarnitine derived from glucose metabolism, is of particular interest as it may reflect the regulatory role of acetyl-CoA on substrate switching and metabolic flexibility ^23^. Its levels have been shown to increase in insulin-related conditions ^24^. Conversely, elevated valine levels were observed in individuals with lower muscle strength, and our analyses indicated significant enrichment in the branched-chain amino acids (BCAA)/aromatic amino acid (AAA) pathways (see **Table 5**, **Figure 5**). Elevated levels of BCAAs and AAAs are recognized as reliable markers of early metabolic dysregulation and play a key role in muscle metabolism, even in older adults ^25^. Previous studies have demonstrated that BCAA levels were inversely associated with sarcopenia in a cohort of 189 older community-dwelling individuals^26^. Further, Aleman-Mateo and colleagues found that BCAAs were negatively associated with sarcopenia while being positively correlated with muscle mass ^27^. However, these findings have not been consistently confirmed in other studies, where BCAA levels were lower in older, sarcopenic community-dwelling individuals ^28^. Interestingly, our data reveals a relationship between early markers of insulin resistance and muscle strength, without any direct association with muscle mass, as already reported ^29^.

We also identified that the inositol-phosphate metabolism was significantly associated with muscle strength. Myo-inositol emerged as a central metabolite, with higher levels observed in individuals with reduced handgrip performance. While the role of myo-inositol in various metabolic diseases has been well-documented, its involvement in muscle metabolism, especially in relation to muscle mass and strength, remains less understood. However, recent studies have linked elevated circulating myo-inositol to conditions associated with reduced muscle strength, such as cachexia and muscle wasting ^30^. Interestingly, both *in vitro* and *in vivo* experiments have shown that reducing the accumulation of myo-inositol in muscle cells can prevent muscle wasting, which is thought to occur through a mechanism involving the inhibition of inositol monophosphatase activity ^31^. These findings suggest that myo-inositol may play a previously underappreciated role in muscle metabolism and could be involved in the regulation of muscle strength and mass.

In women, only two metabolites — glycerol and dimethylsulfone — were identified as being positively associated with muscle mass and function. Elevated circulating glycerol levels are often linked to lipolysis and conditions where insulin action is impaired ^32^. However, the relationship between circulating glycerol levels and muscle mass is not well-established, making it difficult to explain, based on our current data, why individuals with higher lean mass exhibit increased glycerol levels. Dimethylsulfone, that can originate from dietary sources or be produced from methionine through specific microbial activity, was positively associated with muscle strength in women from our study and may suggest early alterations in methionine metabolism. Methionine and its metabolites play a crucial role in combating oxidative stress, acting as radical scavengers and supporting glutathione production. Reduced methionine levels have also been reported in frail individuals with low mobility ^33^. Our findings on dimethylsulfone warrant further investigation to determine whether it could serve as an early marker or proxy for methionine metabolism related to muscle health in older people.

### A low nutritional status is related to reduced insulin sensitivity but not protein intake

Malnutrition is common among older individuals, and its consequences extend beyond physical effects, impacting clinical outcomes, disease recovery, and increasing morbidity and mortality rates ^34^. It also contributes to involuntary weight loss and muscle mass loss, heightening the risk of sarcopenia, a condition closely associated with frailty ^35^. In this study, we identified 22 metabolites that were associated with malnutrition (MNA), a factor often closely associated with frailty. Notably, five metabolites were linked to BCAA metabolism, as supported by significant pathway enrichment analyses (**Table 5**, **Figure 5**), showing higher circulating levels in malnourished individuals. While other studies have reported reduced BCAA levels in older individuals with malnutrition, often linked to poor protein intake and sarcopenia ^36^, this was not observed in our cohort. In our study, none of the identified metabolites were associated with protein or leucine intake, nor with lean mass. Instead, high circulating levels of BCAAs and their metabolites are often linked to compromised insulin sensitivity. This is further reinforced by the presence of alpha-hydroxybutyrate, another metabolite related to amino acid metabolism and recognized as an early marker of insulin resistance ^37^, which was negatively associated with nutritional status in our study. In addition, malnutrition is known to impair insulin sensitivity, and its prevalence is higher among individuals with Type 2 Diabetes (T2D) ^38^. Interestingly, higher HOMA-IR levels were significantly associated with frailty in older individuals over 65 in the NHANES III cohort ^39^, suggesting that HOMA-IR could serve as a novel risk marker for frailty in insulin-resistant older populations ^29^. Moreover, anabolic resistance, which is associated with malnutrition regardless of caloric intake, has been linked to low muscle mass proposed as a hallmark of poor nutritional status ^40^.

### Limits and strengths

Our study has several strengths. First, the associations between the metabolome and other variables were examined in a large, well-characterized cohort, benefiting from comprehensive phenotyping of frailty status. Additionally, in the study population sex was well balanced, which enabled sex-stratified analyses with large sample sizes.

Our study also has several limitations. The overall above-average health and low prevalence and incidence of frailty in our study population could be a reason for the lack of a strong association between frailty-related factors and the metabolome. Another important factor is the choice of methods to measure the metabolome. While NMR offers robustness and reliability for large cohorts like BASE-II, combining it with other methods such as LC-MS, which excels in less concentrated to trace biomarker discovery, could enhance the detection of key metabolites and yield deeper insights into the metabolomic signature of frailty.

## Conclusions

In conclusion, our metabolomics analysis of the BASE-II cohort revealed that, at least in this rather healthy population, the metabolomic fingerprint of frailty was weak. Nevertheless, variables closely associated with frailty, such as muscle strength and nutritional status, were associated with several metabolites from the NMR signatures. This included amino acids involved in BCAA and AAA catabolism, as well as metabolites like sugars and alpha-hydroxybutyrate, connected to muscle strength and nutritional condition, respectively. Interestingly, few associations between muscle mass and the metabolome were found. However, the common signature was primarily composed of early markers of impaired insulin sensitivity, which suggests that in our study population early signs of insulin resistance could be associated with future impairments with respect to muscle health. This association possibly is mediated through diminished anabolic efficiency. Although the sample size and the modest impact of these factors on the metabolomic profiles limited the predictive power of our analyses, further studies on this cohort could help clarify the complex interplay between mobility, nutrition, and metabolism in the frailty syndrome.

## Supporting information

p values

p values

## Data Availability

All data produced in the present study are available upon reasonable request to the authors

## Acknowledgments

The project was funded by iSITE Clermont CAP2025. This article uses data and samples from the Berlin Aging Study II (BASE-II). BASE-II was supported by the German Federal Ministry of Education and Research under grant numbers #01UW0808; #16SV5536K, #16SV5537, #16SV5538, #16SV5837, #01GL1716A, and #01GL1716B.

## Statement of authors’ contributions to manuscript

CB, DD, ID, KN and SP designed research; CC and LD performed RMN analyses; CB, EM, NV and RS analyzed data and performed statistical analysis; CB wrote the first draft with major inputs of ID and SP and critical comments from YB, KN, NV, RS and VMV.

ID and SP are joint last authors and had primary responsibility for final content; All authors have read and approved the final manuscript.

## References

1. Collard RM, Boter H, Schoevers RA, Oude Voshaar RC. Prevalence of Frailty in Community-Dwelling Older Persons: A Systematic Review. J Am Geriatr Soc 2012;60:1487–92.

2. Abellan van Kan G, Rolland Y, Bergman H, Morley JE, Kritchevsky SB, Vellas B. The I.A.N.A Task Force on frailty assessment of older people in clinical practice. J Nutr Health Aging 2008;12:29–37.

3. Fried LP, Tangen CM, Walston J, Newman AB, Hirsch C, Gottdiener J et al. Frailty in older adults: evidence for a phenotype. J Gerontol Biol Sci Med Sci 2001;56.

4. Clegg A, Young J, Iliffe S, Rikkert MO, Rockwood K. Frailty in elderly people. The Lancet 2013;381:752–62.

5. Rodríguez-Mañas L, Féart C, Mann G, Viña J, Chatterji S, Chodzko-Zajko W et al. Searching for an operational definition of frailty: a Delphi method based consensus statement: the frailty operative definition-consensus conference project. J Gerontol A Biol Sci Med Sci 2013;68:62– 67.

6. Theou O, Brothers TD, Mitnitski A, Rockwood K. Operationalization of Frailty Using Eight Commonly Used Scales and Comparison of Their Ability to Predict All-Cause Mortality. J Am Geriatr Soc 2013;61:1537–51.

7. Fernandez-Garrido J, Ruiz-Ros V, Buigues C, Navarro-Martinez R, Cauli O. Clinical features of prefrail older individuals and emerging peripheral biomarkers: a systematic review. Arch Gerontol Geriatr 2014;59:7–17.

8. Pujos-Guillot E, Pétéra M, Jacquemin J, Centeno D, Lyan B, Montoliu I et al. Identification of Pre-frailty Sub-Phenotypes in Elderly Using Metabolomics. 2019 doi:10.3389/fphys.2018.01903.

9. Demuth I, Banszerus V, Drewelies J, Düzel S, Seeland U, Spira D et al. Cohort profile: follow-up of a Berlin Aging Study II (BASE-II) subsample as part of the GendAge study. BMJ Open 2021;11:e045576.

10. Spira D, Buchmann N, Nikolov J, Demuth I, Steinhagen-Thiessen E, Eckardt R et al. Association of Low Lean Mass With Frailty and Physical Performance: A Comparison Between Two Operational Definitions of Sarcopenia—Data From the Berlin Aging Study II (BASE-II). J Gerontol A Biol Sci Med Sci 2015;70:779–784.

11. Orme JG, Reis J, Herz EJ. Factorial and discriminant validity of the center for epidemiological studies depression (CES-D) scale. J Clin Psychol 1986;42:28–33.

12. Podsiadlo D, Richardson S. The Timed “Up & Go”: A Test of Basic Functional Mobility for Frail Elderly Persons. J Am Geriatr Soc 1991;39:142–8.

13. Vetter VM, Kalies CH, Sommerer Y, Spira D, Drewelies J, Regitz-Zagrosek V et al. Relationship Between 5 Epigenetic Clocks, Telomere Length, and Functional Capacity Assessed in Older Adults: Cross-Sectional and Longitudinal Analyses. J Gerontol Ser A 2022;77:1724–1733.

14. Vellas B, Guigoz Y, Garry PJ, Nourhashemi F, Bennahum D, Lauque S et al. The mini nutritional assessment (MNA) and its use in grading the nutritional state of elderly patients. Nutrition 1999;15:116–22.

15. Lefort G, Liaubet L, Canlet C, Tardivel P, Père M-C, Quesnel H et al. ASICS: an R package for a whole analysis workflow of 1D 1H NMR spectra. Bioinformatics 2019;35:4356–63.

16. R Core Team. R: A Language and Environment for Statistical Computing. 2022 https://www.R-project.org/.

17. Lefort G, Liaubet L, Marty-Gasset N, Canlet C, Vialaneix N, Servien R. Joint Automatic Metabolite Identification and Quantification of a Set of ^1^ H NMR Spectra. Anal Chem 2021;93:2861–70.

18. Vetter VM, Drewelies J, Düzel S, Homann J, Meyer-Arndt L, Braun J et al. Change in body weight of older adults before and during the COVID-19 pandemic: longitudinal results from the Berlin Aging Study II. J Nutr Health Aging 2024;28:100206.

19. Pan Y, Li Y, Liu P, Zhang Y, Li B, Liu Z et al. Metabolomics-Based Frailty Biomarkers in Older Chinese Adults. 2022 doi:10.3389/fmed.2021.830723.

20. Brunelli L, Davin A, Sestito G, Mimmi MC, Simone G, Balducci C et al. Plasmatic Hippuric Acid as a Hallmark of Frailty in an Italian Cohort: The Mediation Effect of Fruit–Vegetable Intake. J Gerontol Ser A 2021;76:2081–9.

21. Sepulveda M, Arauna D, Garcia F, Albala C, Palomo I, Fuentes E. Frailty in Aging and the Search for the Optimal Biomarker: A Review. Biomedicines 2022;10.

22. Lopez-Pedrosa JM, Camprubi-Robles M, Guzman-Rolo G, Lopez-Gonzalez A, Garcia-Almeida JM, Sanz-Paris A et al. The Vicious Cycle of Type 2 Diabetes Mellitus and Skeletal Muscle Atrophy: Clinical. Biochem Nutr Bases 2024;16.

23. Schooneman MG, Vaz FM, Houten SM, Soeters MR. Acylcarnitines: reflecting or inflicting insulin resistance? Diabetes 2013;62:1–8.

24. Tsintzas K, Chokkalingam K, Jewell K, Norton L, Macdonald IA, Constantin-Teodosiu D. Elevated free fatty acids attenuate the insulin-induced suppression of PDK4 gene expression in human skeletal muscle: potential role of intramuscular long-chain acyl-coenzyme A. J Clin Endocrinol Metab 2007;92:3967–72.

25. Meng L, Yang R, Wang D, Wu W, Shi J, Shen J et al. Specific lysophosphatidylcholine and acylcarnitine related to sarcopenia and its components in older men. BMC Geriatr 2022;22.

26. Lu Y, Karagounis LG, Ng TP, Carre C, Narang V, Wong G et al. Systemic and Metabolic Signature of Sarcopenia in Community-Dwelling Older Adults. J Gerontol Biol Sci Med Sci 2020;75:309–17.

27. Aleman-Mateo H, Macias L, Esparza-Romero J, Astiazaran-Garcia H, Blancas AL. Physiological effects beyond the significant gain in muscle mass in sarcopenic elderly men: evidence from a randomized clinical trial using a protein-rich food. Clin Interv Aging 2012;7:225–34.

28. Picca A, Calvani R, Cesari M, Landi F, Bernabei R, Coelho-Junior HJ et al. Biomarkers of Physical Frailty and Sarcopenia: Coming up to the Place? Int J Mol Sci 2020;21.

29. Barzilay JI, Cotsonis GA, Walston J, Schwartz AV, Satterfield S, Miljkovic I et al. Insulin resistance is associated with decreased quadriceps muscle strength in nondiabetic adults aged >or=70 years. Diabetes Care 2009;32:736–738.

30. Yang Q-J, Zhao J-R, Hao J, Li B, Huo Y, Han Y-L et al. Serum and urine metabolomics study reveals a distinct diagnostic model for cancer cachexia. 2018 doi:10.1002/jcsm.12246.

31. Lee JH, Kim HJ, Kim SW, Um J, Jung DW, Williams DR. Inhibited inositol monophosphatase and decreased myo-inositol concentration improve wasting in skeletal muscles. Clin Transl Med 2020;10.

32. Mahendran Y, Cederberg H, Vangipurapu J, Kangas AJ, Soininen P, Kuusisto J et al. Glycerol and fatty acids in serum predict the development of hyperglycemia and type 2 diabetes in Finnish men. Diabetes Care 2013;36:3732–8.

33. Kameda M, Teruya T, Yanagida M, Kondoh H. Frailty markers comprise blood metabolites involved in antioxidation, cognition, and mobility. Proc Natl Acad Sci U A 2020;117:9483–9.

34. Norman K, Haß U, Pirlich M. Malnutrition in Older Adults—Recent Advances and Remaining Challenges. Nutrients 2021;13:2764.

35. Ni Lochlainn M, Cox NJ, Wilson T, Hayhoe RPG, Ramsay SE, Granic A et al. Nutrition and Frailty: Opportunities for Prevention and Treatment. Nutrients 2021;13:2349.

36. Ter Borg S, Luiking YC, van Helvoort A, Boirie Y, Schols JMGA, de Groot CPGM. Low Levels of Branched Chain Amino Acids, Eicosapentaenoic Acid and Micronutrients Are Associated with Low Muscle Mass, Strength and Function in Community-Dwelling Older Adults. J Nutr Health Aging 2019;23:27–34.

37. Gall WE, Beebe K, Lawton KA, Adam KP, Mitchell MW, Nakhle PJ et al. alpha-hydroxybutyrate is an early biomarker of insulin resistance and glucose intolerance in a nondiabetic population. PLoS ONE 2010;5.

38. Vischer UM, Perrenoud L, Genet C, Ardigo S, Registe-Rameau Y, Herrmann FR. The high prevalence of malnutrition in elderly diabetic patients: implications for anti-diabetic drug treatments. Diabet Med 2010;27:918–24.

39. Peng P-S, Kao T-W, Chang P-K, Chen W-L, Peng P-J, Wu L-W. Association between HOMA-IR and Frailty among U.S. Middle-Aged Elder Popul Sci Rep 2019;9.

40. Jensen GL, Cederholm T, Correia M, Gonzalez MC, Fukushima R, Higashiguchi T et al. GLIM Criteria for the Diagnosis of Malnutrition: A Consensus Report From the Global Clinical Nutrition Community. JPEN J Parenter Enter Nutr 2019;43:32–40.

