## Supplementary methods and figures for "Metabolomic signatures associated with frailty, muscle strength and nutritional status in a cohort of older people"

Suppementary methods:

Data quality control:

For metabolomic data, serum samples at baseline were used. Three spectra containing abnormal peaks in the water region were removed from the analysis. Thus, a total of 920 subjects, with both quantified and clinical data, were available for analysis (**Figure 1**). Principal Component Analysis (PCA) (R package FactoMineR [28], version 2.4) was used to explore the obtained quantifications, identify potential outliers and visualize the effects of experimental and clinical variables. The projection of the 920 samples of the BASE-II visually highlighted 19 atypical samples (**Supplementary Figure 1**) with a total quantification > 0.8 or < 0.1 (**Supplementary Figure 2**). These atypical samples were found systematically associated with poor overall reconstructions of the original spectrum by ASICS (not shown) and were then removed from the analyses, leading to a sample size of 901 (see flowchart in **Figure 1**). For the 901 individual’s dataset, no missing values were reported in metabolomics or frailty score measures. Missing values on the other variables are reported in the figures and tables legends when applied.

Differential analyses with linear models

Numerical variables were described independently using the average and standard deviation, and categorical variables using the number of subjects per modality and percentage. For each variable, a comparison between T0 and T1 was made using statistical tests such as: paired Student's *t*-test for average comparisons [29], McNemar's chi-square test for proportion comparisons [30,31], or McNemar’s test with continuity correction when validity conditions were not met.

Identification of metabolic pathways associated with frailty in a cohort of older people - Supplementary information

*Table 1- List of variables used in the analyses of the BASE-II cohort and their definition.*

| Variable name | Definition | Type | Levels (if relevant) or [min ; max] |
|---|---|---|---|
| Sex | Subject sex | Categorical with 2 levels | Man /Woman |
| Body Mass Index (BMI) | Body Mass Index (kg/m²) | Numeric | [17.69 ; 47.68] at T0<br>[17.17 ; 38.27] at T1 |
| AgeDiff | Duration of follow-up corresponding to the difference between age at the end of follow-up (T1) and age at the recruitment (T0) (in years) | Numeric | [4.39 ; 10.37] |
| Age | Subject age (in years) | Numeric | [60.16 ; 84.63] at T0<br>[64.91 ; 94.07] at T1 |
| Fried frailty score | Score of Fried (points) | Numerical | [0 ; 3] at T0<br>[0 ; 4] at T1 |
| Fried frailty index | Frailty category based on Fried's score | Categorical with 3 levels,<br>but a recoding of the variable with 2 levels was used in the analyses | No frail: score = 0 point<br>Pre-fail: score = 1 or 2 points<br>Frail: score = 3 – 5 points<br>The analyses used as No frail vs "pre-frail or frail" |
| Fried frailty unintentional weight loss | Unintentional weight loss, one of the five components of Fried's frailty index | Categorical with 2 levels | Yes: loss of at least 5% of the body weight in the last year<br>No: no loss of at least 5% of the body weight in the last year |

| Variable name | Definition | Type | Levels (if relevant) or [min ; max] |
|---|---|---|---|
| Fried frailty exhaustion | Self-reported exhaustion (one of the five components of Fried's frailty index) was identified based on two questions from the Center for Epidemiological Studies Depression (CES-D) scale (depressive symptomatology questionnaire). | Categorical with 2 levels | Not exhausted/ exhausted |
| Fried frailty weakness | Hand grip strength categories, one of the five components of Fried's frailty index | Categorical with 2 levels | The cut-off values stratified by sex and BMI were used<br>Not weak / weak |
| Fried frailty walking speed | Walking speed (one of the five components of Fried's frailty index) was judged by using the timed Stand Up and Go test. | Categorical with 2 levels | Slow: time < 10 second<br>Not slow: time ≥ 10 second |
| Fried frailty physical activity | Physical activity (one of the five components of Fried's frailty index) was assessed based on how the subjects feel about their level of activity. | Categorical with 2 levels | Low: rarely or never active<br>Not low: active |
| Weight | Weight of the subjects (kilograms), the value associated with Fried's frailty unintentional weight loss | Numeric | [44.80 ; 131.50] at T0<br>[45.01 ; 133.30] at T7 |
| Weight evolution | Percentage of the evolution of the weight between T0 and T1 | Numeric | [-32.94 ; 43.38] |
| Weight absolute Variation | Absolut variation of the weight between T0 and T1 | Numeric | [-28.59 ; 21.69] |
| CES-D score | Center for Epidemiological Studies depression scale (points), the value associated with Fried's frailty exhaustion | Numeric | [4.00 ; 32.00] at T0<br>[0.00 ; 31.00] at T7 |
| CES-D evolution | Percentage of the evolution of the CES-D between T0 and T1 | Numeric | [-100.00 ; 340.00] |

| Variable name | Definition | Type | Levels (if relevant) or [min ; max] |
|---|---|---|---|
| CES-D absolute Variation | Absolut variation of the CES-D between T0 and T1 | Numeric | [-19.00 ; 17.00] |
| Grip strength | Hand grip strength (in kg), the value associated with Fried's frailty weakness | Numeric | [15.00 ; 64.50] at T0<br>[8.50 ; 54.50] at T7 |
| Grip strength evolution | Percentage of the evolution of the hand grip strength between T0 and T1 | Numeric | [-60.32 ; 37.14] |
| Grip strength absolute Variation | Absolut variation of the hand grip strength between T0 and T1 | Numeric | [-31.50 ; 13.00] |
| RAPA score | Rapid Assessment of Physical Activity (RAPA) score (points) | Numeric | [1 ; 7] at T0 and at T7 |
| RAPA evolution | Percentage of the evolution of the RAPA score between T0 and T1 | Numeric | [-83.33 ; 500.00] |
| RAPA absolute Variation | Absolut variation of the RAPA score between T0 and T1 | Numeric | [-5.00 ; 5.00] |
| Fried frailty evolution | Evolution of Fried's frailty index categories between baseline (T0) and the end of follow-up (T1) (see Figure 2) | Categorical with 4 levels | Control: no frail<br>Improve: Frailty enhancement (ex: frail -> pre-frail, see Figure 2)<br>Stable: No change<br>Damage: Frailty degradation (ex: no frail -> pre-frail) |
| Fried frailty evolution binary | Fried frailty evolution in binary variable | Categorical with 2 levels | Damage/ control, stable or improve |

| Variable name | Definition | Type | Levels (if relevant) or [min ; max] |
|---|---|---|---|
| ALM | Appendicular lean mass (in kg) | Numeric | [11.76 ; 34.82] at T0<br>[11.14 ; 33.47] at T7 |
| FatMass | fat mass (in kg/ m²) | Numeric | [3.21 ; 19.79] at T0<br>[3.49 ; 16.50] at T7 |
| Leucine intake | Dietary leucine intake (in g/ day) | Numeric | [1.35 ; 21.23] at T0<br>[0.52 ; 28.18] at T7 |
| Protein intake | Dietary proteins intake (in g/day or g/kg/day) | Numeric | [17.94 ; 277.79] at T0 for g/day<br>[6.88 ; 381.33] at T7 for g/day<br>[0.26 ; 3.19] at T0 for g/kg/day<br>[0.09 ; 3.91] at T7 for g/kg/day |
| Total energy intake | Daily energy intake (Kilojoule/day) | Numeric | [1,927 ; 24,685] at T0<br>[935 ; 32,048] at T7 |
| MNA score | Mini Nutritional Assessment score (points) | Numeric | [19.00 ; 30.00] at T0<br>[16.00 ; 30.00] at T7 |

*Table 2- Quantifications# of the NMR spectrum for the whole population (901 subjects of the BASE-II cohort), the men (428 subjects) and the women (478 subjects) of the present study.*

| Metabolites | Mean ± SD<br>N=901 subjects | Mean ± SD<br>N=428 men | Mean ± SD<br>N=478 women |
|---|---|---|---|
| D-Glucose | 0.0665237±0.0348618 | 0.0665087±0.0240275 | 0.0647513±0.0291040 |
| Lactate | 0.0289559±0.0143717 | 0.0296931±0.0107799 | 0.0279420±0.0135337 |
| L-Alanine | 0.0080003±0.0041308 | 0.0078160±0.0022930 | 0.0079200±0.0035278 |
| L-Proline | 0.0068516±0.0034052 | 0.0069672±0.0020378 | 0.0065624±0.0029880 |
| L-Glycine | 0.0063979±0.0035625 | 0.0057700±0.0019399 | 0.0067822±0.0033346 |
| L-Glutamine | 0.0062251±0.0037520 | 0.0058526±0.0026745 | 0.0063291±0.0030035 |
| L-Valine | 0.0041908±0.0021289 | 0.0043748±0.0014325 | 0.0039229±0.0018429 |
| DehydroAscorbicAcid | 0.0041686±0.0020185 | 0.0042164±0.0014878 | 0.0040424±0.0018087 |
| EthylmalonicAcid | 0.0040537±0.0026783 | 0.0041808±0.0020401 | 0.0038884±0.0028325 |
| L-Cystine | 0.0035182±0.0016082 | 0.0033438±0.0009644 | 0.0035976±0.0014226 |
| L-Threonine | 0.0034878±0.0013471 | 0.0035335±0.0011384 | 0.0034271±0.0012940 |
| Taurine | 0.0029650±0.0015267 | 0.0029134±0.0009585 | 0.0029235±0.0012790 |
| D-GlucuronicAcid | 0.0026706±0.0014012 | 0.0026495±0.0009465 | 0.0026086±0.0011382 |
| D-Fucose | 0.0026082±0.0014425 | 0.0025661±0.0009767 | 0.0025943±0.0014248 |
| L-GlutamicAcid | 0.0024959±0.0012424 | 0.0025011±0.0008250 | 0.0024337±0.0010735 |
| L-Serine | 0.0024439±0.0013435 | 0.0023016±0.0009456 | 0.0025147±0.0012030 |
| D-Fructose | 0.0023349±0.0010988 | 0.0022790±0.0007672 | 0.0023382±0.0009821 |
| Levoglucosan | 0.0021770±0.0009397 | 0.0021941±0.0006499 | 0.0021215±0.0008258 |
| 2-HydroxybutyricAcid | 0.0021223±0.0019059 | 0.0023802±0.0017532 | 0.0018943±0.0019762 |
| SebacicAcid | 0.0020944±0.0016910 | 0.0022666±0.0014729 | 0.0019269±0.0017740 |
| L-Leucine | 0.0019280±0.0009394 | 0.0021437±0.0007244 | 0.0016975±0.0007371 |
| L-Isoleucine | 0.0017185±0.0007932 | 0.0018032±0.0005411 | 0.0016128±0.0007158 |

| Metabolites | Mean ± SD<br>N=901 subjects | Mean ± SD<br>N=428 men | Mean ± SD<br>N=478 women |
|---|---|---|---|
| Azelaic Acid | 0.0016658±0.0012650 | 0.0017807±0.0010686 | 0.0015574±0.0013560 |
| 2-Propanol | 0.0016104±0.0030559 | 0.0014667±0.0021645 | 0.0017800±0.0037202 |
| GuanidinoaceticAcid | 0.0016054±0.0007740 | 0.0015939±0.0004821 | 0.0015788±0.0007004 |
| Glycerol | 0.0015596±0.0014483 | 0.0011011±0.0008886 | 0.0019293±0.0014372 |
| 2-Oxoisovalerate | 0.0014695±0.0007989 | 0.0013937±0.0005264 | 0.0015125±0.0008084 |
| beta-HydroxyisovalericAcid | 0.0014211±0.0010585 | 0.0015031±0.0008364 | 0.0013328±0.0011377 |
| D-Mannose | 0.0014038±0.0009466 | 0.0014144±0.0006308 | 0.0013312±0.0007709 |
| PyroglutamicAcid | 0.0013385±0.0006673 | 0.0013135±0.0003829 | 0.0013259±0.0005928 |
| L-Ornithine | 0.0013144±0.0006807 | 0.0013262±0.0004321 | 0.0012714±0.0006215 |
| 2-Oxoglutarate | 0.0013080±0.0005765 | 0.0013326±0.0003969 | 0.0012731±0.0005780 |
| L-Aspartate | 0.0012917±0.0006521 | 0.0012993±0.0004403 | 0.0012549±0.0005766 |
| L-Arginine | 0.0012872±0.0008769 | 0.0012197±0.0005863 | 0.0012836±0.0006855 |
| N-Acetyl-L-Aspartic Acid | 0.0012330±0.0007084 | 0.0011949±0.0003987 | 0.0012189±0.0005624 |
| 3-Hydroxybutyrate | 0.0012197±0.0010854 | 0.0012035±0.0011182 | 0.0012501±0.0010130 |
| Threitol | 0.0011905±0.0006171 | 0.0011789±0.0005829 | 0.0011814±0.0004749 |
| N-Acetylglycine | 0.0011616±0.0004863 | 0.0011327±0.0003166 | 0.0011708±0.0004338 |
| Methanol | 0.0011032±0.0005042 | 0.0011257±0.0004618 | 0.0010892±0.0004939 |
| S-Acetamidomethylcysteine | 0.0010982±0.0005637 | 0.0010404±0.0003098 | 0.0011183±0.0004919 |
| Galactitol | 0.0010735±0.0005993 | 0.0010937±0.0003952 | 0.0010190±0.0004802 |
| Myo-Inositol | 0.0010639±0.0005505 | 0.0010656±0.0003680 | 0.0010336±0.0004708 |
| Argininosuccinic Acid | 0.0010585±0.0005766 | 0.0010717±0.0003517 | 0.0010122±0.0004867 |
| L-Phenylalanine | 0.0010539±0.0004321 | 0.0010574±0.0002469 | 0.0010334±0.0003216 |
| Hypotaurine | 0.0010145±0.0005473 | 0.0009910±0.0003763 | 0.0010137±0.0004861 |
| L-Lysine | 0.0010086±0.0008430 | 0.0008168±0.0006320 | 0.0011346±0.0007333 |

| Metabolites | Mean ± SD<br>N=901 subjects | Mean ± SD<br>N=428 men | Mean ± SD<br>N=478 women |
|---|---|---|---|
| L-Arabitol | 0.0010035±0.0004795 | 0.0009806±0.0005159 | 0.0010218±0.0003709 |
| Phosphocholine | 0.0009828±0.0005037 | 0.0008763±0.0003061 | 0.0010597±0.0004716 |
| Glyceric Acid | 0.0009422±0.0008923 | 0.0009198±0.0005504 | 0.0008836±0.0006024 |
| 3-Methylxanthine | 0.0009309±0.0005049 | 0.0009192±0.0003389 | 0.0009130±0.0004079 |
| Methylmalonic Acid | 0.0009274±0.0005270 | 0.0008867±0.0003612 | 0.0009459±0.0005242 |
| L-Methionine | 0.0009028±0.0003583 | 0.0009187±0.0002293 | 0.0008773±0.0003301 |
| 4-EthylPhenol | 0.0008954±0.0007297 | 0.0009211±0.0009761 | 0.0008826±0.0003971 |
| Creatinine | 0.0008677±0.0003135 | 0.0009630±0.0002538 | 0.0007848±0.0002913 |
| Isovaleric Acid | 0.0008558±0.0006037 | 0.0009170±0.0005619 | 0.0007967±0.0005999 |
| Lactose | 0.0008140±0.0004410 | 0.0008186±0.0002633 | 0.0007832±0.0003634 |
| Betaine | 0.0008065±0.0004513 | 0.0008534±0.0003958 | 0.0007443±0.0003558 |
| Valerate | 0.0007904±0.0005675 | 0.0008360±0.0004320 | 0.0007377±0.0006018 |
| Dihydrothymine | 0.0007871±0.0004527 | 0.0007940±0.0003980 | 0.0007617±0.0003617 |
| Glycogen | 0.0007815±0.0003550 | 0.0007675±0.0002740 | 0.0007742±0.0002549 |
| L-Carnitine | 0.0007707±0.0003235 | 0.0007845±0.0002386 | 0.0007474±0.0002881 |
| CholineChloride | 0.0007377±0.0003537 | 0.0007160±0.0002197 | 0.0007424±0.0003332 |
| D-Maltose | 0.0007339±0.0005011 | 0.0007300±0.0002913 | 0.0007028±0.0004196 |
| 7-Methylxanthine | 0.0007034±0.0002492 | 0.0007090±0.0002183 | 0.0006937±0.0002285 |
| L-Tyrosine | 0.0006885±0.0003465 | 0.0006827±0.0002241 | 0.0006811±0.0003393 |
| D-Sorbitol | 0.0006847±0.0005303 | 0.0007276±0.0005373 | 0.0006219±0.0003550 |
| Citrate | 0.0006767±0.0004025 | 0.0006413±0.0001896 | 0.0006853±0.0003180 |
| Pantothenic Acid | 0.0006390±0.0004146 | 0.0006785±0.0003380 | 0.0005921±0.0004101 |
| GABA | 0.0005655±0.0002613 | 0.0005707±0.0001630 | 0.0005467±0.0002199 |
| 1,3-Diaminopropane | 0.0005587±0.0002777 | 0.0005570±0.0001737 | 0.0005456±0.0002430 |

| Metabolites | Mean ± SD<br>N=901 subjects | Mean ± SD<br>N=428 men | Mean ± SD<br>N=478 women |
|---|---|---|---|
| Malic Acid | 0.0004521±0.0002491 | 0.0004273±0.0002314 | 0.0004715±0.0002121 |
| Acetic Acid | 0.0004461±0.0004323 | 0.0004363±0.0004686 | 0.0004553±0.0003566 |
| 3-MethyladipicAcid | 0.0004193±0.0003477 | 0.0004532±0.0003442 | 0.0003881±0.0003364 |
| Beta-Alanine | 0.0004178±0.0002067 | 0.0004140±0.0001263 | 0.0004113±0.0001878 |
| D-Galactose | 0.0004082±0.0003286 | 0.0003897±0.0002719 | 0.0003993±0.0001923 |
| alpha-Hydroxyisobutyric Acid | 0.0003609±0.0002614 | 0.0003890±0.0002241 | 0.0003317±0.0002671 |
| TMAO | 0.0003498±0.0002416 | 0.0003452±0.0002225 | 0.0003522±0.0002362 |
| O-Acetyl-L-Carnitine | 0.0003124±0.0001711 | 0.0003126±0.0001372 | 0.0003073±0.0001623 |
| Acetoacetate | 0.0002853±0.0002141 | 0.0002744±0.0001857 | 0.0002944±0.0002166 |
| Propylene Glycol | 0.0002778±0.0001530 | 0.0002774±0.0001100 | 0.0002694±0.0001237 |
| 3-PhenylPropionic Acid | 0.0002371±0.0001188 | 0.0002349±0.0000688 | 0.0002329±0.0001067 |
| Dimethylsulfone | 0.0001831±0.0006026 | 0.0001550±0.0001200 | 0.0001880±0.0006747 |

### As said in Section 2.3, obtained quantification are relative quantifications (see (34) for details), which value cannot be interpreted directly (the value 1 corresponding to the area under the curve of the whole spectrum), but which can be used for comparison between samples and metabolites.

Table 3 Excel_pValues

This file contains a sheet for each outcome tested, and in each sheet, the *p*-values for each metabolite (in rows) for the whole population, for men and women (in columns).

Table 4 Excel_parameters_estimate

This file contains a sheet for each significant outcome for men, for which there is more than 1 metabolite, the estimated parameter of the linear regression model, his standard deviation, and confidence interval at 95% (in columns) for each significant metabolite (in rows).

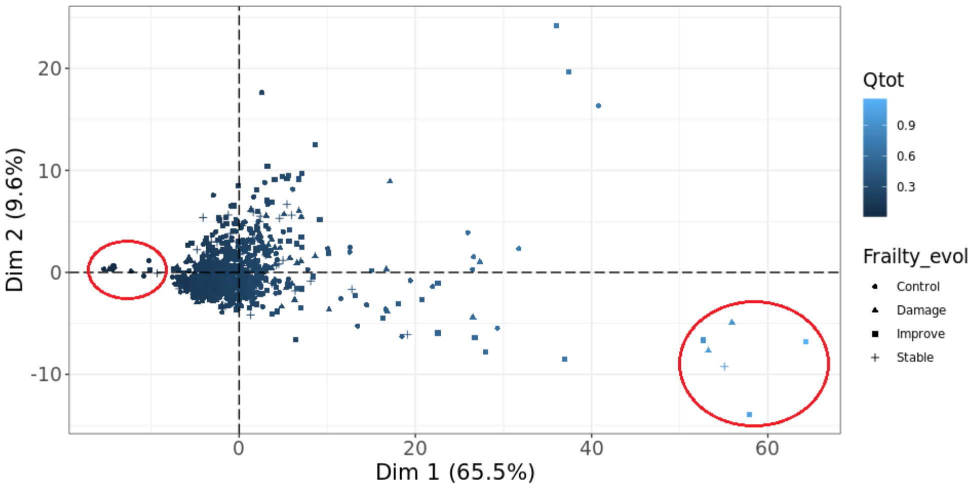


*Figure 1- Projection of subjects obtained by Principal Component Analysis (PCA) on the first two PCs. This analysis was conducted on metabolite quantifications for 920 subjects. The red circles highlighted 19 atypical subjects associated with large and small values of the sum of all identified metabolite concentrations (named Qtot).*

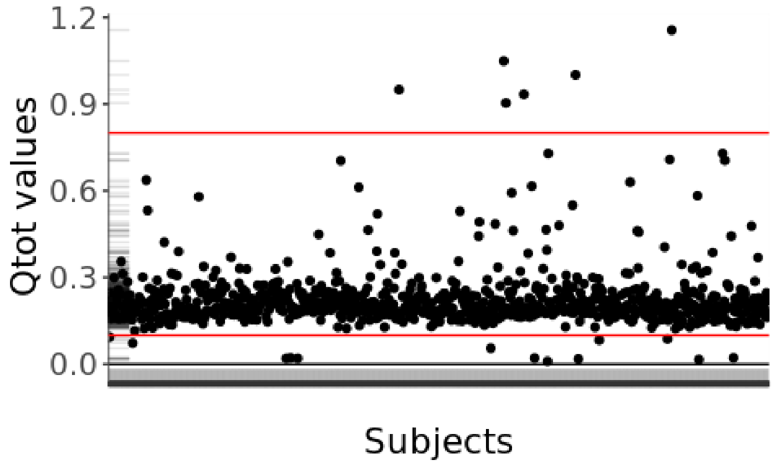


*Figure 2- Dot plot of the sum of all identified metabolite concentrations (named Qtot) according to the subjects of the BASE-II cohort (N=920 subjects, positioned evenly and randomly on the x-axis). The red lines identify the thresholds for atypical subjects systematically associated with poor overall reconstructions of the original spectrum by ASICS (6 subjects for Qtot>0.8 and 13 for Qtot<0.1).*

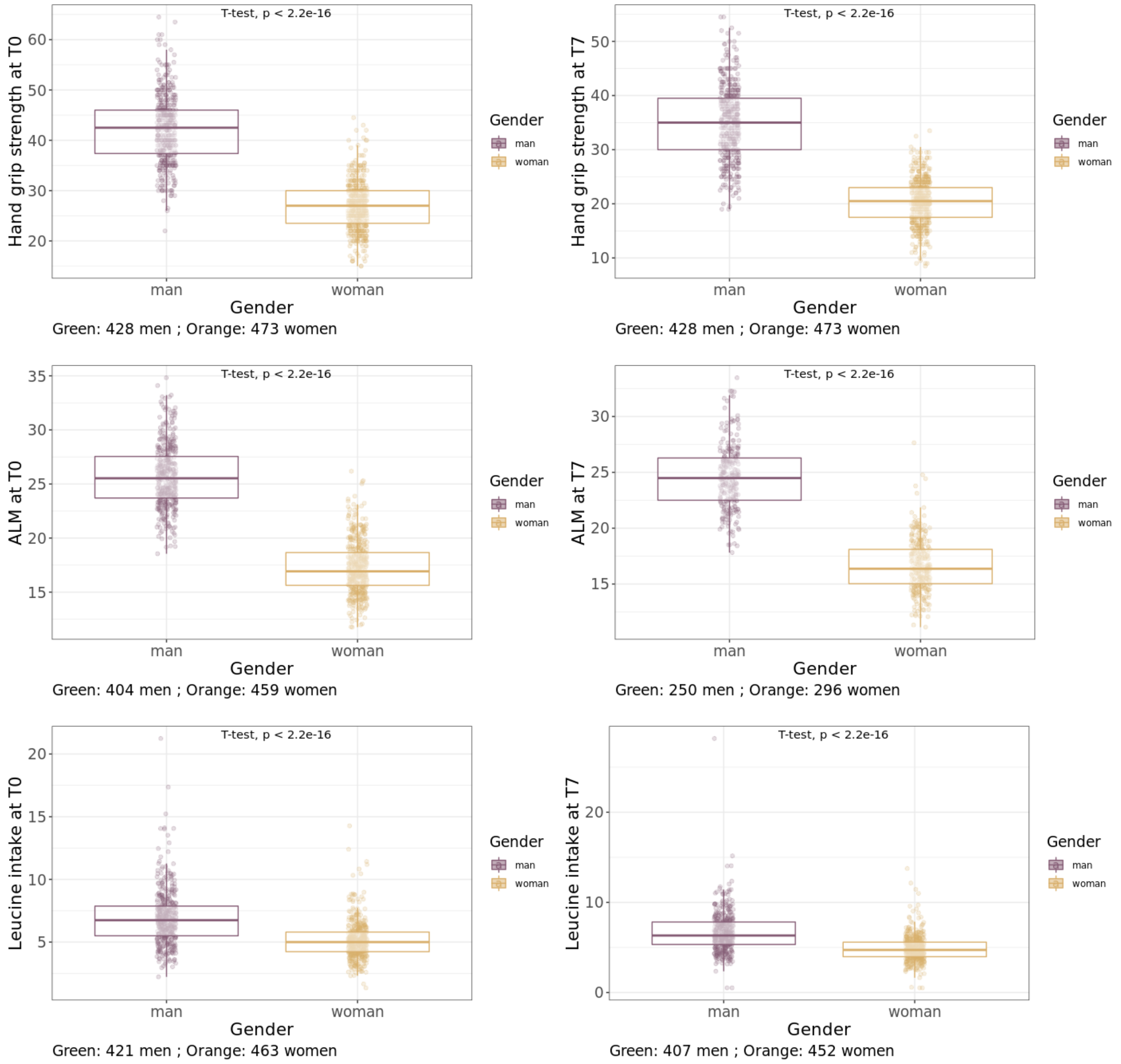


*Figure 3- Boxplots of three examples of clinical variables for which we observe a clear and important sex effect with very low p-value which supports a sex-stratification of the models. The colors were associated with the subjects' sex (purple for men and light orange for women).*

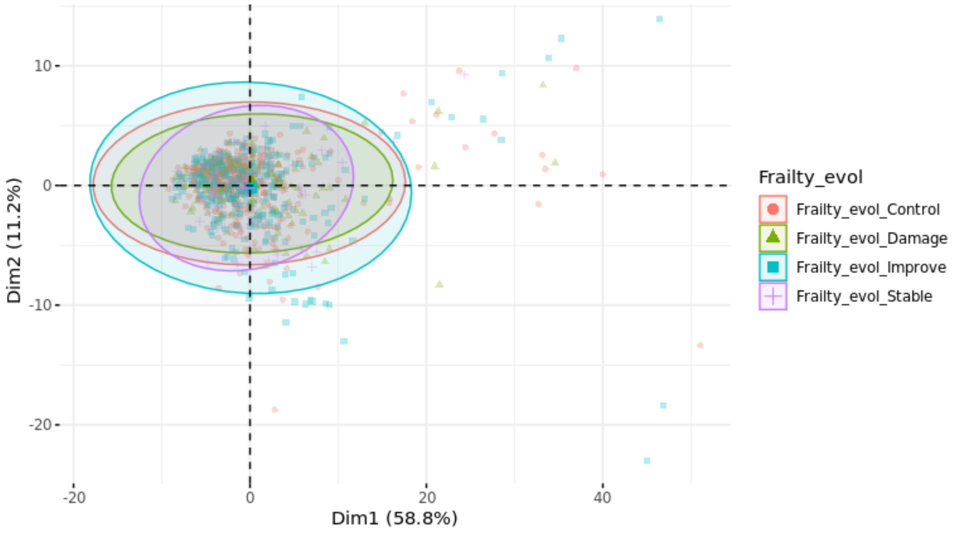


*Figure 4- Principal Component Analysis of metabolite quantifications from serum samples collected at baseline for the 901 subjects in the BASE-II cohort. The different groups of subjects formed by the modalities of Fried's frailty index (on the top) were represented by ellipses of different colors (red for Control, purple for Stable, blue for Improve and green for Damage). All ellipses were superposed, meaning that no group could be distinguished from the others, and that the frailty signal carried by these variables is weak.*

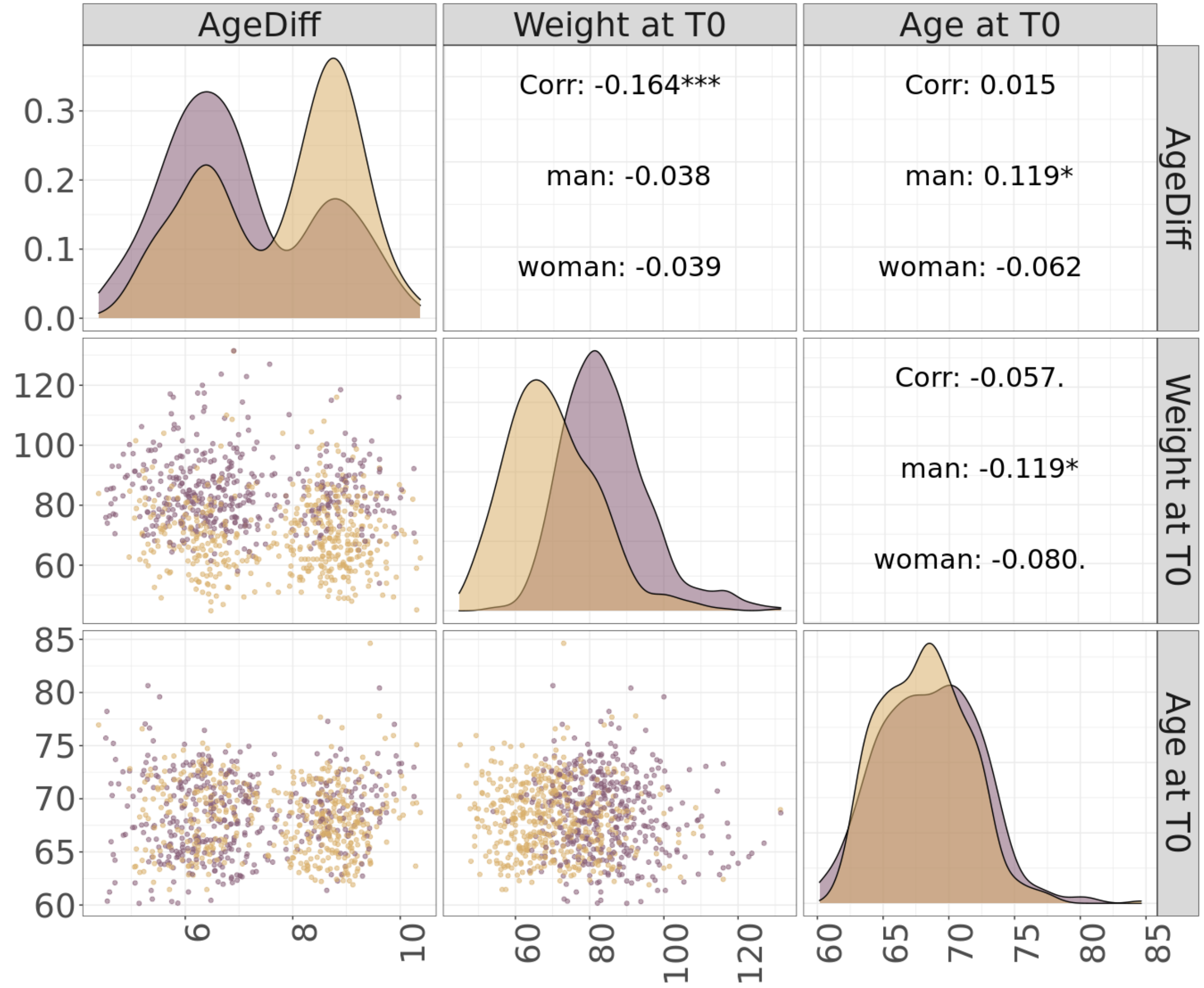


*Figure 5. Scatterplots of each pair of the three variables used as covariates in the linear models (left side). On the right side of the plots, Pearson correlation value and significance are displayed (indicate "*" if the p-value of the correlation test is < 0.001;"" if the p-value is < 0.01; "*" if the p-value is < 0.05; "." if the p-value is < 0.10 and "" otherwise). On the diagonal, the distribution of the variables is colored by sex (light orange for women and purple for men).*
